# Evolving patterns of prevalence and management of hypertension phenotypes in Mexico: A two-decade analysis of nationally representative surveys

**DOI:** 10.1101/2025.08.04.25333004

**Authors:** Carlos A. Fermín-Martínez, Alejandra Núñez-Luna, Enrique C. Guerra, Daniel Ramírez-García, Jeronimo Perezalonso-Espinosa, Kathya Paola Zarco-Morales, Mariana León-Álvarez, Camila Ponce-Acosta, Natalia Sofía De la Maza-Bustindui, Arsenio Vargas-Vázquez, Neftali Eduardo Antonio-Villa, Omar Yaxmehen Bello-Chavolla

## Abstract

**BACKGROUND:** Systemic arterial hypertension is a public health concern, and timely diagnosis and management are critical to mitigate long-term effects. Here, we evaluated prevalence trends and determinants of hypertension and its phenotypes in Mexican population over the last two decades.

**METHODS:** We analyzed cross-sectional Mexican Health and Nutrition Surveys (2000-2023), including 141,668 adults aged ≥20 years. Hypertension was defined as self-reported diagnosis or blood pressure (BP) ≥140/90 mmHg; undiagnosed hypertension (UDH) as BP ≥140/90 mmHg without prior diagnosis; and untreated hypertension (UTH) as prior diagnosis without treatment. UDH was classified as isolated systolic (ISH), isolated diastolic (IDH), or systolic-diastolic hypertension (SDH). We assessed prevalence trends with Poisson models, and determinants of UDH and UTH with logistic models.

**RESULTS:** We observed an overall decrease in hypertension prevalence from 2000 to 2023, driven by increases in diagnosed (12.3% to 19.3%) and decreases in undiagnosed (20.7% to 11%) hypertension. IDH and SDH declined over time, while ISH increased, particularly among older adults. Among diagnosed cases, UTH decreased (31% to 19.5%) and BP control improved (40.5% to 69.5%). Despite these trends, by 2023 approximately 8.2 million Mexican adults still had UDH (36.3% of all hypertension cases), 2.8 million remained untreated, and 4.4 million uncontrolled. Lack of diagnosis and treatment were more likely among men, individuals with unhealthy lifestyles, and social disadvantage.

**CONCLUSIONS:** Results highlight evolving trends in hypertension diagnosis, treatment, and control in Mexico, with persistent challenges in UDH and UTH. Strengthening screening, treatment access, and equity is crucial to reduce hypertension-related cardiovascular risk.

## INTRODUCTION

Systemic arterial hypertension is a modifiable risk factor for cardiovascular disease (CVD) that greatly contributes to the global burden of disease and mortality. It remains a major public health concern worldwide, and its prevalence in The Americas has increased by up to 41% over the last three decades^1^. Despite global efforts to improve blood pressure (BP) screening and hypertension treatment, over 40% of women and 50% of men with hypertension remain unaware of their condition, and management remains suboptimal across most low- and middle-income countries ^2,3^. These challenges are driven not only by high rates of unawareness—often linked to sociodemographic inequalities—but also by the underlying pathophysiological complexity of hypertension^2^. The condition may begin with isolated or combined elevations in systolic and diastolic BP, reflecting distinct clinical phenotypes, such as isolated systolic hypertension (ISH), isolated diastolic hypertension (IDH), and systolic-diastolic hypertension (SDH)^4^. These phenotypes not only influence survival but also the risk of complications, requiring tailored management to improve prognosis^5,6^. Meanwhile, untreated arterial hypertension (UTH) is also associated with increased risk of complications, impaired quality of life, and escalating healthcare costs, thereby imposing a burden on healthcare systems, particularly in countries like Mexico^7^. In Mexico, around 40-60% of individuals with hypertension are unaware of their condition, depending on the definition used, and only a third achieved optimal BP control among those diagnosed^8,9^. To date, there is a lack of characterization of the patterns of hypertension phenotypes, as well as the impact of treatment over time in Mexico. An integrated approach would enable the evaluation of our healthcare system’s effectiveness, enhance screening and treatment initiation, and support BP goal achievement over time, providing a clearer understanding of the current state of hypertension in Mexico. In this work, we aimed to characterize the trends in the prevalence of hypertension—both diagnosed and undiagnosed—and its phenotypes in Mexican population using data from over two-decades of the Mexican National Health and Nutrition Surveys (ENSANUT). Moreover, we aimed to identify sociodemographic and clinical factors that influence hypertension awareness and treatment.

## METHODS

### Study Design

We performed a serial cross-sectional analysis utilizing data extracted from ENSANUT spanning the years 2000, 2006, 2012, 2016, 2018, 2020, 2021, 2022, and 2023^10–15^. ENSANUT is representative at a national and regional level, as it uses a multi-stage probabilistic stratified cluster sampling design based on basic geostatistical areas (primary sampling units), localities, households, and individuals. Sample weights are further corrected using the expected response rate obtained from the response rate observed on the previous cycle and calibrated using a post-stratification correction factor to account for over- or underrepresentation of specific subgroups^15^. A detailed outline of each survey design is provided in **Supplementary Table 1**. Participants engaged in a comprehensive questionnaire capturing demographic, socioeconomic, and health-related information, along with a physical examination involving BP and anthropometric measurements. Additionally, a random subsample from each cycle underwent a supplementary biochemical assessment, including serum samples for fasting glucose, lipid profile, and glycated hemoglobin (HbA1c).

### Hypertension and High-normal BP

Hypertension was defined as a self-reported prior diagnosis, the use of BP-lowering medications, a systolic BP (SBP) ≥140 mmHg, or a diastolic BP (DBP) ≥90 mmHg (using the average of at least two different measurements). High-normal BP was defined as either a SBP of 130-139 mmHg or a DBP of 85-89 mmHg. These definitions are based on guidelines of the European Society of Cardiology (ESC) and the European Society of Hypertension (ESH)^16,17^. Undiagnosed hypertension (UDH) was defined in individuals without prior self-reported diagnosis who otherwise fulfilled the definition of hypertension. As a sensitivity analysis, we contrasted ESC/ESH definitions with those based on the American College of Cardiology/American Heart Association (ACC/AHA) guidelines^17,18^, which define elevated BP as SBP 120-129 mmHg, and hypertension as SBP ≥130 mmHg or DBP ≥80 mmHg.

### Hypertension Phenotypes

Patients with UDH were further classified into additional subgroups using the ESC/ESH cutoffs including ISH, IDH and SDH ^19,20^. They were defined as:

- ISH: SBP ≥140 mmHg with DBP <90 mmHg.
- IDH: SBP <140 mmHg with DBP ≥90 mmHg.
- SDH: SBP ≥140 mmHg with DBP ≥90 mmHg.

Among participants with diagnosed hypertension, UTH was defined as those who did not report receiving treatment in the initial interview; BP medication assessment was not available for ENSANUT 2020^14^. Finally, BP goal achievement among participants with diagnosed hypertension was defined as either a SBP <130 mmHg for participants aged <65 years or <140mmHg for those aged ≥65 years and/or DBP <80 mmHg regardless of age, as per ESC/ESH guidelines^16^.

### Covariates

Diabetes was defined by either self-report (previous diagnosis or using insulin or oral diabetes medications), fasting plasma glucose ≥126 mg/dL or HbA1c ≥6.5%. CVD was defined as a self-reported prior diagnosis of either myocardial infarction, heart failure or stroke. We used body mass index (BMI) to categorize participants with normal weight (18.5-24.9 kg/m^2^), overweight (25-29.9 kg/m^2^) or obesity (≥30 kg/m^2^). Smoking status was categorized as never, former, and current smokers, while alcohol consumption was divided in non-drinkers, occasional drinkers, and daily drinkers. Education level was classified as No Education, Primary (Elementary school), Secondary (High-school or technician) and Tertiary Education (college or above). We also used information about social security affiliation and area of residence (rural and urban). Participants who responded “yes” to the question “Do you speak an indigenous language?” were considered to have an indigenous identity. To assess state-level social disadvantage, we used the density-independent social lag index (DISLI), derived from the residuals of a linear model regressing the social lag index (SLI) onto population density. The SLI is a composite measure of access to education, health care, dwelling quality, and basic services in Mexico^21,22^. DISLI is available for the years 2000, 2005, 2010, 2015, and 2020, and each measurement was matched to the nearest ENSANUT cycle by year. DISLI values were categorized into quartiles and labeled as very low, low, middle, and high.

### Statistical Analyses

The prevalence of hypertension, high-normal BP, and hypertension phenotypes was estimated among participants with complete BP measurements using ENSANUT sample weights with the *survey* R package^23^. We also conducted weighted subgroup analyses to examine prevalence trends stratified by age (20-39, 40-59, or ≥60 years old), sex, BMI categories (obesity vs. no obesity), and diabetes status. To assess trends in prevalence, we used weighted Poisson regression models with ENSANUT year as the predictor, estimating prevalence ratios (PR) and 95% confidence intervals (95% CI) to represent the annual change in prevalence.

Among participants with hypertension, we assessed potential factors associated with UDH and UTH using weighted logistic regression models. We tested the following predictors: age (10-year increments), sex, smoking status, alcohol intake, prior diabetes diagnosis, BMI-defined obesity, education level, lack of social security affiliation, indigenous identity, high DISLI and ENSANUT year. All statistical analyses were conducted using R software version 4.5.1 and p-values thresholds are estimated for a two-sided significance level of α = 0.05.

## RESULTS

### Study Population

Among 245,519 adults aged ≥20 years who completed a health interview in ENSANUT between 2000 and 2023, we identified 141,668 individuals with complete BP measurements. These participants were included in the analysis to estimate the prevalence of hypertension in Mexico. Additionally, we identified 36,158 adults with hypertension (either previously diagnosed or newly identified) and 16,411 adults with a prior diagnosis of hypertension and complete covariate data. These subgroups were used to examine predictors of undiagnosed and untreated hypertension, respectively. A detailed flowchart of participant selection is presented in **Supplementary Figure 1**.

Across all ENSANUT cycles, participants with hypertension and high-normal BP were older than those with normal BP, with the highest median age seen in patients with ISH, followed by those with diagnosed hypertension and SDH. Most hypertension categories showed a male predominance over the years, except for normal BP and diagnosed hypertension, where women accounted for over 60% of the population. Participants reporting never smoking and not currently drinking were most common among those with normal BP and diagnosed hypertension. Regarding socioeconomic factors, higher education was more frequent in the normal BP group, while indigenous identity was less common among those with diagnosed hypertension. Lack of social security and high DISLI were prevalent across most hypertension categories but consistently lower in diagnosed hypertension. Overall, diagnosed hypertension was linked to comparatively better socioeconomic conditions, with moderate improvements over time. BMI was highest in participants with SDH, followed by diagnosed hypertension, IDH, high-normal BP, and normal BP, with ISH having the lowest BMI. WHtR, a marker of abdominal adiposity, showed a similar pattern, except for individuals with normal BP having lower WHtR than those with ISH. The prevalence of diabetes, CKD, and CVD was highest among those with diagnosed hypertension, followed by ISH, and lowest in participants with normal BP (**Supplementary Tables 2-10**).

### Trends in the Prevalence of Hypertension in Mexico

The prevalence of overall hypertension had a slight decrease over time (PR 0.996, 95%CI 0.995-0.998), ranging from 33.1% (32.2-34.0%) in 2000 to 30.3% (27.5-33.1%) in 2023. This was attributable to a decrease in the prevalence of UDH over time (PR 0.970, 95%CI 0.968-0.972), changing from 20.7% (19.8-21.6%) in 2000 to 11.0% (9.2-12.8%) in 2023, with a corresponding increase in the prevalence of diagnosed hypertension (PR 1.016, 95%CI 1.014-1.017), from 12.3% (11.7-12.9) in the year 2000 to 19.3% (16.8-21.8%) in 2023 (**Figure 1, Supplementary Table 11**). By the year 2023, we estimate that 63.7% of participants with hypertension were aware of their condition, representing a total of 14,409,647 adults (11,410,841-17,408,453); however, unawareness was still 36.3%, which represents a total of 8,220,799 adults (6,336,363-10,105,236) with undiagnosed hypertension in Mexico. Similar trends were observed using the ACC/AHA definitions, although with higher prevalences of both total and undiagnosed hypertension (**Supplementary Table 12**).

**Figure 1.**
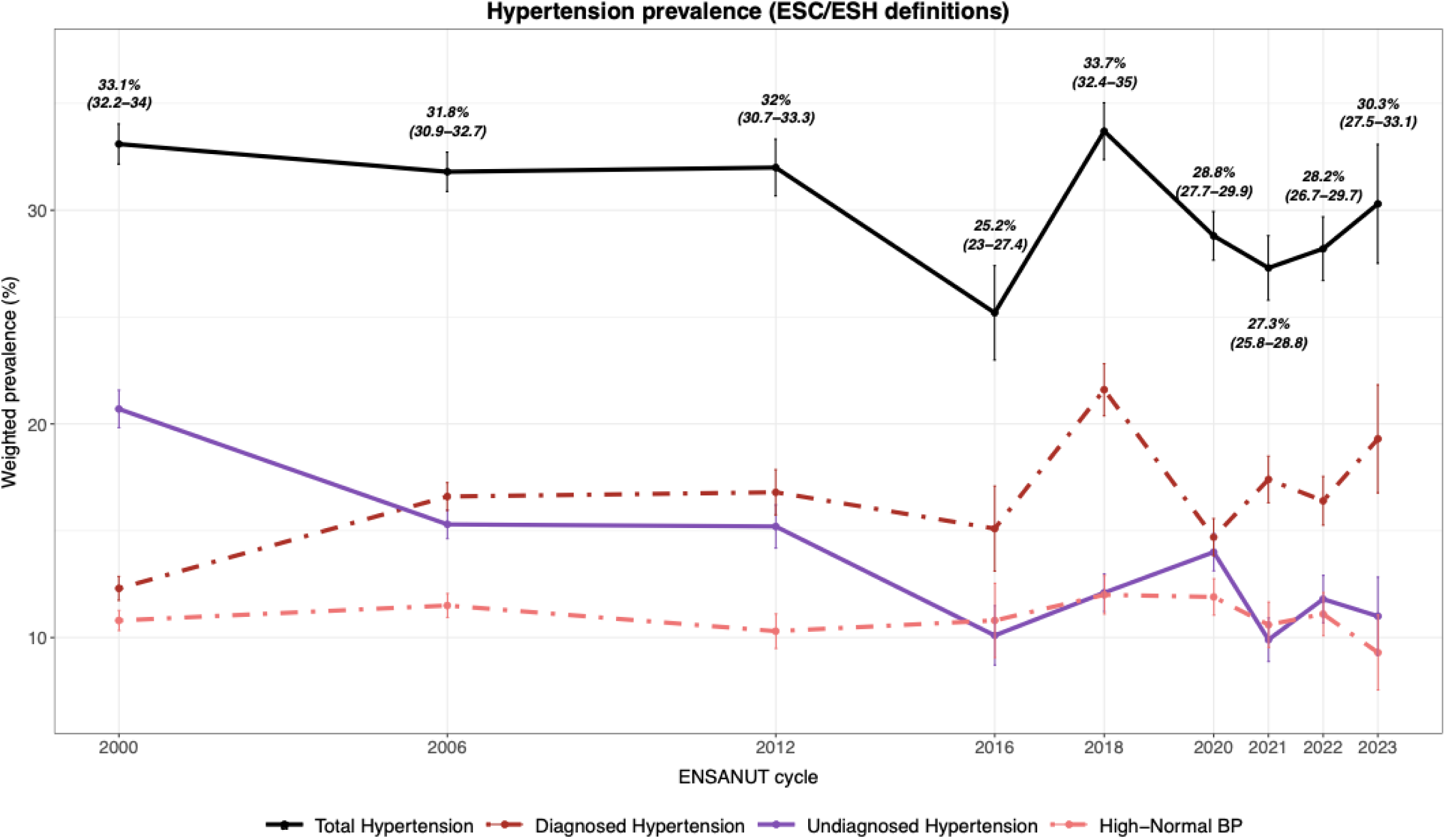
Hypertension prevalence trends in Mexico, 2000-2023. Weighted prevalence of diagnosed, undiagnosed and total systemic arterial hypertension, and high-normal blood pressure across ENSANUT cycles (2000-2023). Estimates are based on the European Society of Cardiology/European Society of Hypertension (ESC/ESH) definitions of hypertension and include 95% confidence intervals.

### Trends in the Prevalence of UDH Phenotypes in Mexico

Among UDH phenotypes, the prevalence of ISH increased significantly over time (PR 1.037, 95%CI 1.034-1.040), rising from 2.4% (2.2-2.6%) in 2000 to 5.4% (4.4-6.4%) in 2023. In contrast, IDH declined markedly (PR 0.922, 95%CI 0.919-0.925), dropping from 12.5% (11.8-13.2%) to 2.0% (1.1-2.9%) over the same period. SDH also decreased (PR 0.970, 95%CI 0.967-0.973), from 5.8% (5.4-6.1%) to 3.6% (2.6-4.6%) (**Figure 2A, Supplementary Figure 3**). The observed decrease in UDH prevalence over time appears to be largely driven by decreases in IDH, which represented 60.1% of all UDH cases in 2000 but only 18.3% in 2023, and by corresponding increases in ISH, which went from 11.7% of all UDH cases in 2000 to 49.2% by 2023 (**Supplementary Figure 4**).

**Figure 2.**
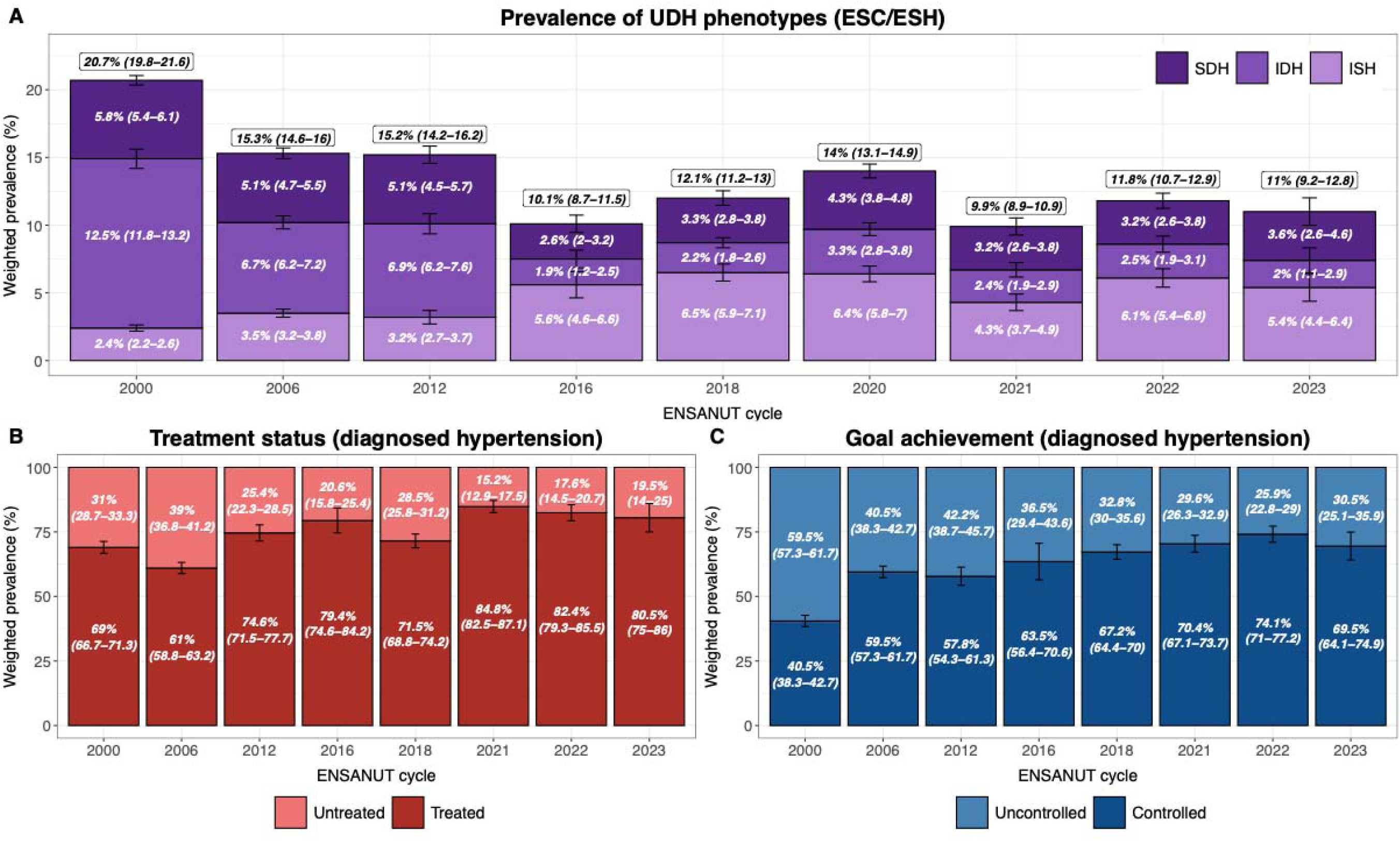
Undiagnosed hypertension phenotypes, treatment, and control in Mexico. **(A)** Prevalence of undiagnosed hypertension (UDH, top label) and its phenotypes: isolated systolic hypertension (ISH), isolated diastolic hypertension (IDH), and systolic-diastolic hypertension (SDH) in the Mexican population. **(B)** Percentage of individuals with previously diagnosed hypertension receiving pharmacological treatment. **(C)** Percentage of individuals with previously diagnosed hypertension achieving blood pressure control goals. Estimates are based on ENSANUT 2000-2023 data and include 95% confidence intervals. Information on hypertension treatment was not collected in 2020.

### Trends in the Prevalence of Diagnosed Hypertension Phenotypes in Mexico

Among participants with previously diagnosed hypertension, we observed that 31% (28.7-33.3%) were not receiving any treatment in 2000; however, this percentage was substantially reduced to 19.5% (14.0-25.0%) by 2023 (**Figure 2B**). Accordingly, we observed a significant increase in the prevalence of BP goal achievement among adults with diagnosed hypertension, which went from 40.5% (38.3-42.7%) in 2000 to 69.5% (64.1-74.9%) in 2023 (**Figure 2C**). By 2023, an estimated 2.8 million adults (2,809,307; 95%CI 1,982,396-3,636,218) with previously diagnosed hypertension remained untreated, and 4.4 million (4,389,092; 3,222,363-5,555,821) had not achieved BP goals. More concerning, nearly one million (905,694; 441,861-1,369,526) were both untreated and had uncontrolled BP levels.

### Modifiers of undiagnosed hypertension prevalence

UDH was most predominant in individuals aged ≥60 years, followed by those aged 40-59, and this pattern was consistent in all cycles (**Figure 3A**). Regarding sex, we observed a higher prevalence of UDH in males compared to females, except in the year 2000 when females were more affected (**Figure 3B**). UDH was also more prevalent in participants with BMI-defined obesity and in those with diabetes (**Figures 2C-D**). ISH prevalence consistently increased over time, particularly for subjects ≥60 years, and remained higher for men and subjects with diabetes (**Supplementary Figure 5**). IDH prevalence decreased over time and was higher in men and in those with obesity; interestingly, it was more prevalent in those aged 40-59 years compared to older adults (**Supplementary Figure 6**). SDH also decreased in prevalence over time, with more pronounced decreases in subjects ≥60 years (**Supplementary Figure 7**).

**Figure 3.**
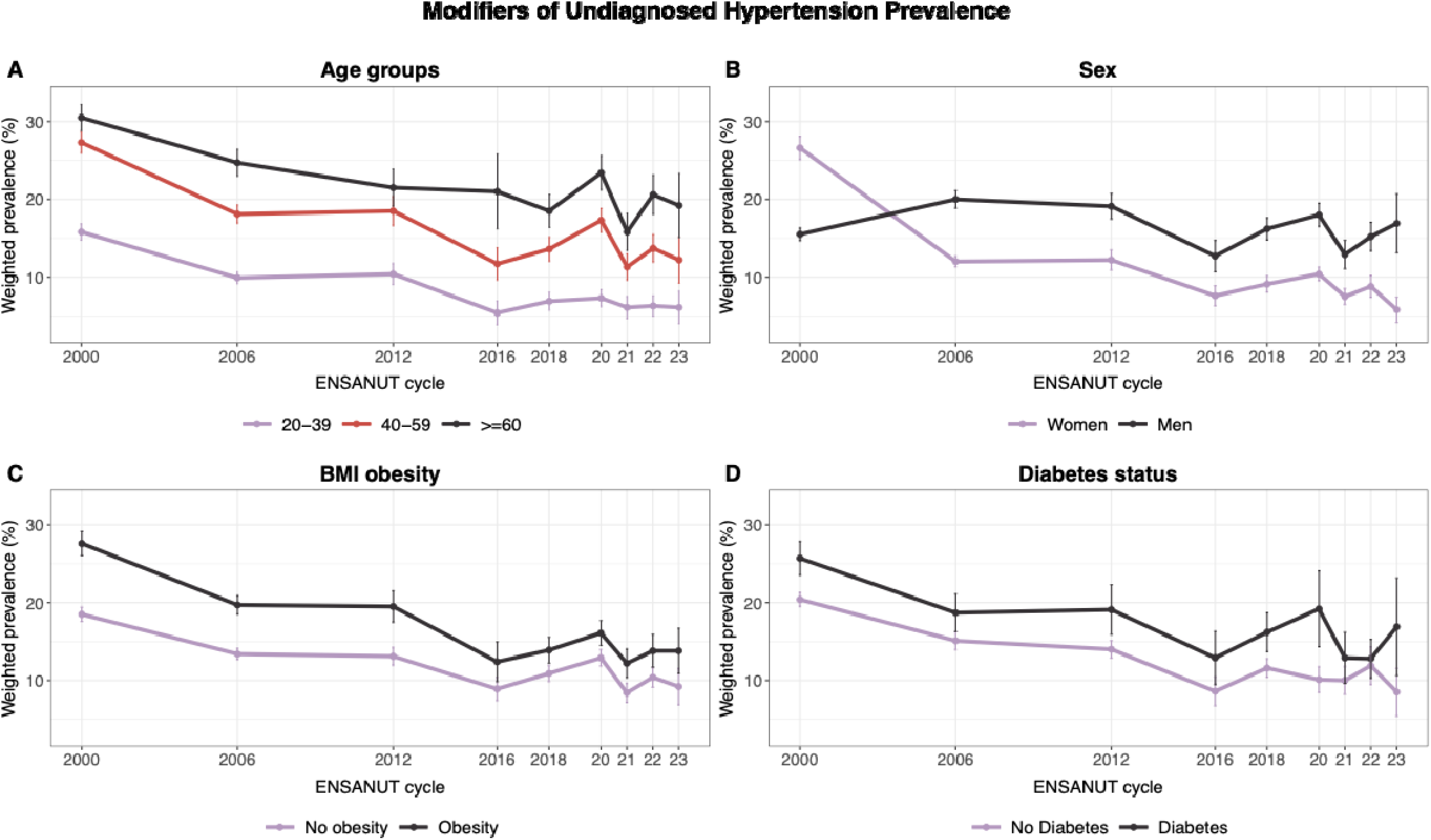
Modifiers of undiagnosed hypertension prevalence in Mexico. Trends in undiagnosed hypertension based on ENSANUT 2000-2023 data, stratified by **(A)** age group (20-39, 40-59, or ≥60 years), **(B)** sex, **(C)** body mass index (BMI)-defined obesity (≥30 kg/m^2^), and **(D)** diabetes status (defined as either prior diagnosis, receiving pharmacological treatment, HbA1c ≥6.5% or fasting plasma glucose ≥126 mg/dL).

### Predictors of untreated and undiagnosed hypertension

Finally, we explored potential determinants of undiagnosed and untreated hypertension using logistic regression models. Higher odds of both lack of diagnosis and lack of treatment were observed among men, former and current smokers (vs. never smokers), occasional and daily drinkers (vs. non-drinkers), individuals identifying as indigenous, those without social security, and those living in areas of high marginalization (high DISLI). In contrast, older adults, individuals with obesity, and those with a prior diabetes diagnosis were more likely to be aware of their condition and to receive treatment. Living in rural areas was associated with higher odds of being undiagnosed, while higher educational attainment was linked to greater likelihood of treatment. Notably, the odds of undiagnosed and untreated hypertension declined over time since 2000 (**Figure 4**).

**Figure 4.**
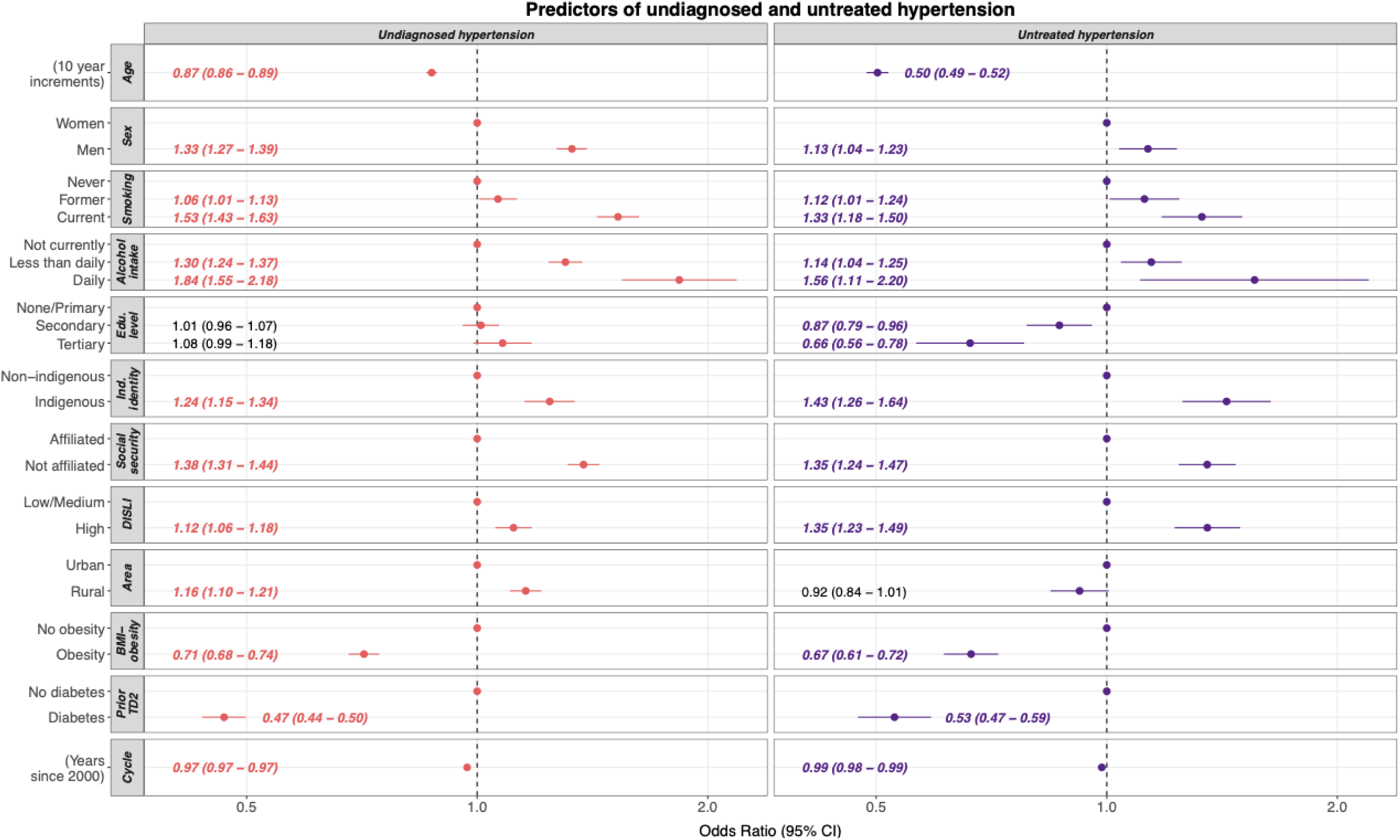
Predictors of undiagnosed and untreated hypertension. Weighted logistic regression models pooling ENSANUT 2000-2023 data, showing odds ratios (OR) with 95% confidence intervals (95% CI) for factors associated with **(A)** undiagnosed hypertension (n = 36,158 adults with overall hypertension) and **(B)** untreated hypertension (n = 16,411 adults with diagnosed hypertension) in Mexico. ENSANUT 2020 was excluded from the untreated analysis due to missing treatment data (see **Supplementary Figure 1**). Predictors included age (per 10-year increments), sex, smoking status, alcohol intake, education level, indigenous identity, social security affiliation, density-independent social lag index (DISLI), area of residence, body mass index (BMI)-defined obesity, prior diagnosis of type 2 diabetes (T2D), and ENSANUT cycle (number of years since 2000).

## DISCUSSION

In this serial cross-sectional study of nationally representative surveys from 2000 to 2023, we characterized trends in hypertension prevalence and its undiagnosed and diagnosed phenotypes in Mexico. Over this 23-year period, hypertension prevalence slightly declined, driven by a marked reduction in UDH and a corresponding rise in diagnosed hypertension. Among UDH cases, prevalence of IDH and SDH decreased, while ISH increased over this period. In those with diagnosed hypertension, treatment rates improved, resulting in BP control for 7 out of 10 adults with hypertension by 2023. Despite this progress, suboptimal diagnosis and management remained high in Mexico in 2023, with an estimated 8.2 million adults (36.3% of all hypertension cases) undiagnosed, 2.8 million (19.5% of those diagnosed) untreated, and 4.4 million (30.5% of those diagnosed) with uncontrolled blood pressure, particularly among men, individuals with unhealthy lifestyles, and those facing sociodemographic disadvantage, highlighting the need to improve detection, treatment, and control in these groups. Although several reports have documented trends in hypertension awareness, treatment, and control at both global and regional levels^3,24,25^, this study represents, to our knowledge, the first long-term assessment of the evolution of hypertension patterns and phenotypes in the Mexican population. These findings may inform future policy efforts to reduce the burden of hypertension in the country.

Although we observed a decline in hypertension prevalence, a substantial proportion of Mexican adults remain undiagnosed. The NCD Risk Factor Collaboration estimated that 32.5% of women and 53.1% of men with hypertension in Mexico were unaware of their condition in 2019^3^. In contrast, an independent study of 5,901 adults from Eastern Mexico reported a lower unawareness rate of 28.3% that same year, with higher rates among men, individuals with obesity or diabetes, and residents of a low-income state^26^. Another study conducted during the COVID-19 pandemic found a 30.9% unawareness rate, which was associated with male sex, older age, low education, physical inactivity, and prior COVID-19 infection^27^. In our 2023 analysis, we estimate that 36.3% of adults with hypertension (∼8.2 million people) were still unaware of their diagnosis. A main concern regarding reports on UDH prevalence are related to how they change over time, especially as countries shift their focus toward increased BP screening^28^. A longitudinal analysis from the SAGE study reported that over a third of adults aged 50 years or older with UDH in Mexico remain undiagnosed after five years, highlighting men, individuals living in rural areas, unmarried, without health insurance, and overweight^7^. Similarly, a longitudinal analysis across four middle-income countries, including Mexico, reported that only around 30% of undiagnosed individuals ≥40 years eventually received a diagnosis (27% in Mexico) after five to nine years, with similar rates observed in China, Indonesia, and South Africa^29^. Our study adds to this evidence, showing that hypertension unawareness remains more prevalent among individuals living in rural and marginalized areas, without health insurance, with low education level, and who self-identify as indigenous. These findings suggest that, while BP screening efforts in Mexico have substantially reduced the prevalence of UDH, there has been little improvement in longstanding disparities in access to screening services over the past two decades.

The assessment of UDH phenotypes in our study provides a more nuanced view of the hypertension landscape in Mexico. In 2023, ISH was the most common phenotype (∼5.4%, accounting for 57% of UDH cases), with increases likely driven by population aging, as ISH predominates in older adults, in whom treatment significantly improves cardiovascular outcomes^30–32^. Although the most pronounced increases occurred in adults ≥60 years, men, and individuals with diabetes, milder increases were also observed in younger adults, where ISH has been linked to sustained SBP elevation, hypertension-mediated organ damage, and higher CVD risk^33,34^. In contrast, IDH showed a six-fold decrease over the past two decades (from 12.5% to 2%), remaining most common in younger adults, a trend also reported in other populations^35,36^. IDH is associated with increased CVD risk, particularly in younger adults and using higher diagnostic thresholds^37–39^. SDH prevalence remained relatively stable, with a slight decline. By 2023, its prevalence was broadly comparable to that of ISH, depending on the definition, a pattern also observed in other populations^40–42^. Recent studies suggest that SDH may confer greater CVD risk than ISH or IDH, especially at higher BP levels^4,40^, consistent with genetic evidence linking elevations in both SBP and DBP to coronary disease and shorter lifespan^43^. The decline in undiagnosed IDH, and to a lesser extent SDH, may reflect improvements in BP screening and earlier detection. Conversely, the rise in ISH likely reflects population aging. While all UDH phenotypes carry cardiovascular risk, ISH holds particularly strong prognostic value, especially in older adults^31,44^. These trends highlight the need to standardize phenotype definitions, enhance awareness of ISH as a high-risk phenotype, and consider hypertension presentation when initiating treatment^6^.

Despite marked increases in hypertension diagnosis, treatment and optimal BP control remain as ongoing challenges to reduce the burden of hypertension worldwide^1,3^. One of the primary drivers of UTH is the lack of diagnosis awareness, which has been estimated to be ∼40% among Mexican adults with hypertension^9^. During the 23-year period observed in our study, UTH decreased from 31% in 2000 to 19.5% in 2023, while BP goal achievement increased from 40.5% in 2000 to 69.5% in 2023. Although these improvements in BP diagnosis, treatment and control in Mexico are encouraging and not substantially behind those observed in some high-income countries^24^, the burden of elevated BP levels remains as one of the main drivers of cardiovascular disease burden in Mexico^45,46^, where persistent disparities in access to screening and treatment continue to pose significant challenges. Our findings show that UDH and UTH were more common among men, individuals who smoked or consumed alcohol regularly, those living in highly marginalized areas, with lower educational attainment, without social security, and among people identifying as Indigenous. In contrast, diagnosis and treatment were more likely among women, older adults, and individuals with obesity or a prior diabetes diagnosis. These patterns highlight the influence of social and behavioral determinants—such as healthcare access, education, Indigenous identity, and lifestyle factors—on hypertension care. They also reflect the frequent coexistence of hypertension with metabolic conditions and high-risk behaviors, which may shape how individuals engage with the healthcare system^47–49^. Targeted policies are needed to improve hypertension diagnosis and management, particularly among socially and economically vulnerable populations.

Our study has several strengths. By leveraging two decades of nationally representative data, we were able to provide a reliable overview of hypertension diagnosis and management in Mexico, enabling the identification of prevalence trends across over time. Furthermore, our study offers the first reports on UDH phenotypes in Mexico, providing valuable insights into hypertension presentation before treatment initiation and identifying groups less likely to be diagnosed during routine screenings. Additionally, by pooling data from over 141,000 Mexican adults, we characterized determinants of UDH and UTH, highlighting populations at higher risk of lack of diagnosis and treatment who likely experience an increased burden of hypertension-related complications. Despite these strengths, several limitations should be acknowledged when interpreting our findings. First, we restricted the analysis of ISH, IDH, and SDH phenotypes to individuals with UDH, as we considered that treatment initiation and lifestyle modifications following diagnosis could substantially alter blood pressure levels, limiting the reliability of phenotype classification in diagnosed cases. Second, the absence of detailed information on the type and adherence to antihypertensive medications across ENSANUT cycles limited our ability to assess changes in treatment strategies and their contribution to BP control over time. Third, analyses of the determinants of UDH and UTH relied on cross-sectional data, which prevents causal inference. Future research should prioritize longitudinal studies to evaluate individual trajectories of hypertension phenotypes and to assess the impact of public health interventions on their management and related outcomes in the Mexican population. Likewise, further investigation is needed to understand treatment adherence and effectiveness across different hypertension phenotypes, and to explore the social and contextual factors influencing the diagnosis, monitoring, and control of hypertension.

In conclusion, this study provides a comprehensive assessment of trends in hypertension diagnosis, treatment, and control in Mexico over the past two decades. While hypertension diagnosis rates have increased over time, UDH remains a pressing public health concern, particularly among individuals with ISH. Moreover, despite notable reductions in the prevalence of UTH, 30% of adults with diagnosed hypertension still fail to meet BP targets as of 2023. Our findings underscore the complexity of hypertension control in Mexico and highlight the need for sustained screening efforts, broader access to effective treatment, and robust monitoring systems. Addressing sociodemographic disparities in access to hypertension care is essential to reduce the burden of elevated BP and achieve equitable cardiovascular health outcomes in vulnerable populations in Mexico.

## ACKNOWLEDGMENTS

This project was registered and approved by the Research Committee at Instituto Nacional de Geriatría, project number DI-PI-009-2024. CAFM, ECG, and JPE are enrolled at the PECEM Program of the Faculty of Medicine at UNAM and are supported by SECIHTI.

## AUTHOR CONTRIBUTIONS

Research idea and study design: CAFM, ANL, ECG, AVV, OYBC; data acquisition and processing: CAFM, ANL, ECG, OYBC; statistical analysis: CAFM, ANL, OYBC; analysis/interpretation: CAFM, ANL, ECG, DRG, NEAV, OYBC; manuscript drafting: CAFM, ANL, ECG, DRG, KPZM, MLA, CPA, NSMB, AVV, NEAV, OYBC; supervision or mentorship: OYBC. Each author contributed important intellectual content during manuscript drafting or revision and accepts accountability for the overall work by ensuring that questions pertaining to the accuracy or integrity of any portion of the work are appropriately investigated and resolved.

## DATA AVAILABILITY

All code and materials regarding ENSANUT are available for reproducibility of results http://github.com/oyaxbell/hypertension_ensanut/.

## ETHICS APPROVAL AND CONSENT TO PARTICIPATE

This project was registered and approved by the Research and Ethics Committee at Instituto Nacional de Geriatría, project number DI-PI-009/2024.

