## Supplementary Material for "Evolving patterns of prevalence and management of hypertension phenotypes in Mexico: A two-decade analysis of nationally representative surveys"

#### SUPPLEMENTARY TABLES

1. Summary of sampling designs for ENSA/ENSANUT surveys....2
2. ENSA 2000 population table....4
3. ENSANUT 2006 population table....6
4. ENSANUT 2012 population table....8
5. ENSANUT 2016 population table.... 10
6. ENSANUT 2018 population table.... 12
7. ENSANUT 2020 population table.... 14
8. ENSANUT 2021 population table.... 16
9. ENSANUT 2022 population table.... 18
10. ENSANUT 2023 population table .... 20
11. ESC/ESH hypertension prevalence from 2000-2023 .... 22
12. ACC/AHA hypertension prevalence from 2000-2023 .... 23
13. ESC/ESH hypertension prevalence by sex .... 24
14. ESC/ESH hypertension prevalence by age group.... 25
15. ESC/ESH hypertension prevalence by obesity status.... 27
16. ESC/ESH hypertension prevalence by diabetes status .... 28

#### SUPPLEMENTARY FIGURES

1. Flowchart of participant selection.... 29
2. ACC/AHA hypertension prevalence trends.... 30
3. Proportions of undiagnosed hypertension phenotypes (ESC/ESH).... 31
4. Proportions of undiagnosed hypertension phenotypes (ESC/ESH).... 32
5. Trends of isolated systolic hypertension (ISH). .... 33
6. Trends of isolated diastolic hypertension (IDH). .... 34
7. Trends of systolic-diastolic hypertension (SDH). .... 35

### SUPPLEMENTARY TABLES

**Supplementary Table 1. Summary of sampling designs for ENSA/ENSANUT surveys**. (Probabilistic, stratified, multi-stage).

<table>
<tr><th>Survey</th><th>Strata</th><th>Primary Sampling Unit (PSU)</th><th>Sample selection</th></tr>
<tr><td>ENSA 2000</td><td>State<br>+<br>Locality size*<br>(Urban, rural)</td><td>Municipality<br>(355 PSUs)</td><td>1) Municipalities (probability proportional to size [PPS] sampling based on the number of households).<br>2) AGEB[†] (PPS sampling).<br>3) Blocks (simple random sampling [SRS]).<br>4) Households (SRS).<br>5) Individuals: for each household 1 school-age child (0-9 years), 1 adolescent (10-19 y), and 1 adult (≥20 y) were selected (SRS).</td></tr>
<tr><td>ENSANUT 2006</td><td>State<br>+<br>Locality size*<br>(Metropolitan, urban, rural)<br>+<br>OPORTUNIDADES program[‡]<br>(Enrolled, not enrolled)</td><td>AGEB[†]<br>(1346 PSUs)<br><br>Oversampling in households enrolled in OPORTUNIDADES</td><td rowspan="3"><u>Urban and metropolitan</u><br><br>1) AGEB[†] (PPS sampling).<br>2) Blocks (PPS sampling).<br>3) Households (systematic sampling).<br>4) Individuals: 1 school-age child, 1 adolescent, 1 adult, and 1 user of health services (SRS).<br><br><u>Rural areas</u><br><br>1) AGEB[†] (PPS sampling).<br>2) Localities (PPS sampling).<br>3) Pseudo-blocks (systematic sampling).<br>4) Groups of ~15 households (SRS).<br>5) Individuals: 1 school-age child, 1 adolescent, 1 adult, and 1 user of health services (SRS).</td></tr>
<tr><td>ENSANUT 2012</td><td>State<br>+<br>Urban stratum<br>(Metropolitan, urban, rural, newly created)<br>+<br>Marginality[§]<br>(High SLI, low SLI)</td><td>AGEB[†]<br>(1592 PSUs)<br><br>Oversampling in AGEBs with high SLI</td></tr>
<tr><td>ENSANUT 2016</td><td>State<br>+<br>Locality size*<br>(Metropolitan, urban complement, rural)<br>+<br>Marginality[§]<br>(High, medium, low SLI)</td><td>AGEB[†]<br>(326 PSUs)<br><br>Oversampling in AGEBs with high SLI</td></tr>
</table>

<table>
<tr>
<td>ENSANUT 2018</td>
<td>State<br>+<br>Locality size*<br>(Metropolitan, urban complement, rural)<br>+<br>Sociodemographic characteristics¶</td>
<td><u>Urban and metropolitan</u><br>Blocks / groups of blocks<br><br><u>Rural areas</u><br>AGEBs / groups of AGEBs<br><br>(3153 PSUs)</td>
<td><u>Urban and metropolitan</u><br>1) PSUs (SRS).<br>2) Households (SRS).<br>3) Individuals: 1 preschooler (0-4 y), 1 school-age child (5-9 y), 1 adolescent (10-19 y), and 1 adult (≥20 y).<br><br><u>Rural areas</u><br>1) Primary sampling units were selected with SRS.<br>2) Groups of ~5 households (SRS).<br>3) Individuals: see above.</td>
</tr>
<tr>
<td>ENSANUT 2020</td>
<td rowspan="4">State<br>+<br>Locality size*<br>(Metropolitan, urban, rural)</td>
<td>AGEB†<br>(427 PSUs)<br>Oversampling in Guanajuato state</td>
<td rowspan="4">Same as 2006-2016 except for the selection of individuals:<br><br>• In 2020 and 2021: 1 preschooler, 1 school-age child, 1 adolescent, 1 adult 20-34 years, 1 adult 35-50 years, 1 adult ≥50 years and 1 user of health services (SRS).<br>• In 2022: 1 preschooler, 1 school-age child, 1 adolescent, 1 adult 20-44 years, 1 adult ≥45 years and up to 2 users of health services (SRS).</td>
</tr>
<tr>
<td>ENSANUT 2021</td>
<td>AGEB†<br>(454 PSUs)<br>Oversampling in Guanajuato state</td>
</tr>
<tr>
<td>ENSANUT 2022</td>
<td>AGEB†<br>(432 PSUs)<br>Oversampling in Guanajuato and Nuevo Leon states</td>
</tr>
<tr>
<td>ENSANUT 2023</td>
<td>AGEB†<br>(260 PSUs)<br>Oversampling in Sinaloa, Sonora, and Guanajuato states</td>
</tr>
</table>

*Localities are categorized in most surveys according to the number of residents in ***metropolitan*** (capital cities or cities/metropolitan areas with ≥100,000 residents), ***urban/urban complement*** (areas with 2,500 – 999,999 residents not included in the metropolitan category), and ***rural*** (areas with <2,500 residents). For 2000, they were only categorized in rural and urban (≥2,500 residents). For 2012, a "***newly created localities***" category was also created to account for discrepancies between the 2005 and 2010 national censuses.

‡OPORTUNIDADES was a social assistance program run by the Mexican government between 2002 and 2014 which aimed to provide education, health, nutritional and financial aid to population living in extreme poverty. In 2014 it was rebranded as the "PROSPERA" program and later substituted in 2020 by the "Free Healthcare and Medication Program".

†***AGEB: Basic Geostatistical Areas***. They represent the fundamental unit of the National Geostatistical Framework, a system proposed by INEGI that divides Mexico's territory into distinct levels of disaggregation. AGEBs are divided into urban and rural according to population density. https://en.www.inegi.org.mx/temas/mg/

§Marginality and social disadvantage were measured using the ***Social Lag Index (SLI)*** in the years 2000, 2005, 2010, 2015 and 2020.

¶For 2018, multiple ***sociodemographic strata*** were created based on 34 indicators which provide information on dwelling (e.g., overcrowding, access to basic services, etc.) and inhabitants (e.g., education level, employment, entitlement to health services, etc.) characteristics.

**Supplementary Table 2. ENSA 2000 population characteristics by hypertension status**. Abbreviations. BP: Blood Pressure; IDH: Isolated Diastolic Hypertension; ISH: Isolated Systolic Hypertension; SDH: Systolic-Diastolic Hypertension; DISLI: Density Independent Social Lag Index; SBP: Systolic Blood Pressure; DBP: Diastolic Blood Pressure; WHtR: Waist-to-height ratio; BMI: Body-mass index; CKD, Chronic Kidney Disease; CVD, Cardiovascular Disease.

| **ENSA 2000 Characteristic** | **Overall** N = 44,181[1] | **Normal BP** N = 23,732[1] | **High-Normal** N = 4,680[1] | **IDH** N = 5,144[1] | **ISH** N = 1,279[1] | **SDH** N = 2,851[1] | **Diagnosed** N = 6,495[1] | **p-value**[2] |
|---|---|---|---|---|---|---|---|---|
| **Age (years)** | 38 (28, 52) | 33 (26, 42) | 42 (31, 55) | 40 (31, 52) | 58 (43, 70) | 53 (42, 64) | 52 (39, 64) | <0.001 |
| **Female (%)** | 13,890 (31%) | 6,481 (27%) | 1,874 (40%) | 2,277 (44%) | 529 (41%) | 1,293 (45%) | 1,436 (22%) | <0.001 |
| **Smoking (%)** | | | | | | | | <0.001 |
| Never | 16,694 (56%) | 9,330 (59%) | 1,632 (50%) | 1,771 (50%) | 439 (47%) | 938 (47%) | 2,584 (58%) | |
| Former | 5,758 (19%) | 2,409 (15%) | 723 (22%) | 745 (21%) | 266 (28%) | 539 (27%) | 1,076 (24%) | |
| Current | 7,471 (25%) | 3,966 (25%) | 897 (28%) | 1,059 (30%) | 233 (25%) | 528 (26%) | 788 (18%) | |
| Missing | 14,258 | 8,027 | 1,428 | 1,569 | 341 | 846 | 2,047 | |
| **Alcohol intake (%)** | | | | | | | | <0.001 |
| Not currently | 28,693 (65%) | 15,347 (65%) | 2,914 (62%) | 3,046 (59%) | 900 (71%) | 1,798 (63%) | 4,688 (72%) | |
| Less than daily | 14,685 (33%) | 8,005 (34%) | 1,671 (36%) | 1,975 (38%) | 342 (27%) | 965 (34%) | 1,727 (27%) | |
| Daily | 743 (1.7%) | 350 (1.5%) | 93 (2.0%) | 113 (2.2%) | 31 (2.4%) | 81 (2.8%) | 75 (1.2%) | |
| Missing | 60 | 30 | 2 | 10 | 6 | 7 | 5 | |
| **Education level (%)** | | | | | | | | <0.001 |
| None | 1,069 (2.7%) | 397 (1.8%) | 136 (3.3%) | 152 (3.3%) | 47 (4.7%) | 141 (6.0%) | 196 (3.5%) | |
| Primary | 20,727 (52%) | 10,131 (46%) | 2,289 (56%) | 2,491 (53%) | 695 (70%) | 1,490 (63%) | 3,631 (64%) | |
| Secondary | 14,995 (38%) | 9,541 (43%) | 1,374 (33%) | 1,681 (36%) | 211 (21%) | 603 (26%) | 1,585 (28%) | |
| Tertiary | 3,023 (7.6%) | 1,927 (8.8%) | 314 (7.6%) | 349 (7.5%) | 41 (4.1%) | 123 (5.2%) | 269 (4.7%) | |
| Missing | 4,367 | 1,736 | 567 | 471 | 285 | 494 | 814 | |
| **Indigenous identity (%)** | 3,576 (8.2%) | 2,039 (8.7%) | 356 (7.7%) | 445 (8.7%) | 127 (10%) | 255 (9.0%) | 354 (5.5%) | <0.001 |
| Missing | 374 | 197 | 50 | 42 | 15 | 26 | 44 | |

| ENSA 2000 Characteristic | Overall N = 44,181[1] | Normal BP N = 23,732[1] | High-Normal N = 4,680[1] | IDH N = 5,144[1] | ISH N = 1,279[1] | SDH N = 2,851[1] | Diagnosed N = 6,495[1] | p-value[2] |
|---|---|---|---|---|---|---|---|---|
| **Social security (%)** | | | | | | | | <0.001 |
| Affiliated | 19,719 (45%) | 10,008 (42%) | 2,130 (46%) | 2,311 (45%) | 545 (43%) | 1,229 (43%) | 3,496 (54%) | |
| Not affiliated | 24,313 (55%) | 13,633 (58%) | 2,541 (54%) | 2,815 (55%) | 728 (57%) | 1,614 (57%) | 2,982 (46%) | |
| Missing | 149 | 91 | 9 | 18 | 6 | 8 | 17 | |
| **High DISLI (%)** | 11,048 (25%) | 6,497 (27%) | 1,041 (22%) | 1,167 (23%) | 319 (25%) | 662 (23%) | 1,362 (21%) | <0.001 |
| **SBP (mmHg)** | 120 (110, 130) | 110 (110, 120) | 130 (125, 130) | 130 (120, 130) | 140 (140, 150) | 145 (140, 153) | 130 (120, 150) | <0.001 |
| **DBP (mmHg)** | 80 (70, 90) | 75 (70, 80) | 80 (80, 85) | 90 (90, 90) | 80 (75, 81) | 95 (90, 100) | 90 (80, 95) | <0.001 |
| **WHtR** | 0.59 (0.53, 0.66) | 0.57 (0.51, 0.63) | 0.61 (0.55, 0.67) | 0.61 (0.55, 0.67) | 0.63 (0.57, 0.69) | 0.64 (0.58, 0.71) | 0.65 (0.59, 0.72) | <0.001 |
| Missing | 2,315 | 1,399 | 230 | 207 | 66 | 125 | 288 | |
| **BMI (kg/m2)** | 26.9 (23.8, 30.5) | 25.7 (23.0, 29.0) | 27.7 (24.6, 30.9) | 27.9 (24.8, 31.4) | 27.3 (24.3, 31.0) | 28.9 (25.5, 32.9) | 29.3 (25.9, 33.0) | <0.001 |
| **BMI-defined obesity (%)** | 12,164 (28%) | 4,598 (20%) | 1,435 (31%) | 1,715 (34%) | 383 (31%) | 1,164 (42%) | 2,869 (45%) | <0.001 |
| Missing | 518 | 234 | 61 | 38 | 30 | 51 | 104 | |
| **Capillary glucose (mg/dL)** | 96 (84, 113) | 93 (82, 106) | 97 (86, 115) | 96 (85, 115) | 106 (90, 132) | 104 (90, 130) | 104 (88, 129) | <0.001 |
| Missing | 543 | 288 | 43 | 66 | 22 | 49 | 75 | |
| **Prior diabetes diagnosis (%)** | 2,954 (6.7%) | 774 (3.3%) | 316 (6.8%) | 338 (6.6%) | 157 (12%) | 296 (10%) | 1,073 (17%) | <0.001 |
| Missing | 49 | 22 | 4 | 8 | 4 | 8 | 3 | |
| **Prior CKD diagnosis (%)** | 314 (0.7%) | 122 (0.5%) | 27 (0.6%) | 36 (0.7%) | 8 (0.6%) | 16 (0.6%) | 105 (1.6%) | <0.001 |
| Missing | 185 | 91 | 23 | 24 | 7 | 15 | 25 | |
| **Prior CVD diagnosis (%)** | *Data not available for this cycle* | | | | | | | |
| Missing | - | - | - | - | - | - | - | - |

[1]Median (IQR); n (%). [2]Kruskal-Wallis rank sum test; Pearson's Chi-squared test

**Supplementary Table 3. ENSANUT 2006 population characteristics by hypertension status**. Abbreviations. BP: Blood Pressure; IDH: Isolated Diastolic Hypertension; ISH: Isolated Systolic Hypertension; SDH: Systolic-Diastolic Hypertension; DISLI: Density Independent Social Lag Index; SBP: Systolic Blood Pressure; DBP: Diastolic Blood Pressure; WHtR: Waist-to-height ratio; BMI: Body-mass index; FPG, Fasting Plasma Glucose; CKD, Chronic Kidney Disease; CVD, Cardiovascular Disease.

| ENSANUT 2006 Characteristic | Overall N = 33,485[1] | Normal BP N = 18,699[1] | High-Normal N = 3,911[1] | IDH N = 2,317[1] | ISH N = 1,314[1] | SDH N = 1,781[1] | Diagnosed N = 5,463[1] | p-value[2] |
|---|---|---|---|---|---|---|---|---|
| **Age (years)** | 40 (31, 53) | 35 (28, 45) | 42 (33, 55) | 40 (32, 50) | 60 (46, 71) | 52 (40, 63) | 52 (39, 65) | <0.001 |
| **Female (%)** | 20,277 (61%) | 11,868 (63%) | 1,934 (49%) | 1,062 (46%) | 697 (53%) | 846 (48%) | 3,870 (71%) | <0.001 |
| **Smoking (%)** | | | | | | | | <0.001 |
| Never | 20,978 (71%) | 11,921 (73%) | 2,349 (68%) | 1,370 (67%) | 812 (70%) | 1,040 (67%) | 3,486 (72%) | |
| Former | 3,371 (11%) | 1,552 (9.5%) | 429 (12%) | 247 (12%) | 173 (15%) | 226 (14%) | 744 (15%) | |
| Current | 5,106 (17%) | 2,941 (18%) | 678 (20%) | 418 (21%) | 182 (16%) | 296 (19%) | 591 (12%) | |
| Missing | 4,030 | 2,285 | 455 | 282 | 147 | 219 | 642 | |
| **Alcohol intake (%)** | | | | | | | | <0.001 |
| Not currently | 23,172 (69%) | 13,015 (70%) | 2,547 (65%) | 1,452 (63%) | 937 (71%) | 1,170 (66%) | 4,051 (74%) | |
| Less than daily | 9,867 (29%) | 5,509 (29%) | 1,291 (33%) | 819 (35%) | 341 (26%) | 564 (32%) | 1,343 (25%) | |
| Daily | 438 (1.3%) | 170 (0.9%) | 70 (1.8%) | 46 (2.0%) | 36 (2.7%) | 47 (2.6%) | 69 (1.3%) | |
| Missing | 8 | 5 | 3 | 0 | 0 | 0 | 0 | |
| **Education level (%)** | | | | | | | | <0.001 |
| None | 3,737 (11%) | 1,549 (8.3%) | 520 (13%) | 217 (9.4%) | 306 (23%) | 314 (18%) | 831 (15%) | |
| Primary | 15,056 (45%) | 7,623 (41%) | 1,801 (46%) | 1,042 (45%) | 682 (52%) | 950 (54%) | 2,958 (54%) | |
| Secondary | 11,944 (36%) | 7,755 (42%) | 1,297 (33%) | 861 (37%) | 264 (20%) | 416 (23%) | 1,351 (25%) | |
| Tertiary | 2,618 (7.8%) | 1,704 (9.1%) | 272 (7.0%) | 187 (8.1%) | 55 (4.2%) | 92 (5.2%) | 308 (5.7%) | |
| Missing | 130 | 68 | 21 | 10 | 7 | 9 | 15 | |
| **Indigenous identity (%)** | 7,016 (21%) | 3,961 (21%) | 834 (21%) | 453 (20%) | 318 (24%) | 379 (21%) | 1,071 (20%) | 0.003 |
| Missing | 154 | 81 | 15 | 22 | 6 | 12 | 18 | |

| ENSANUT 2006 Characteristic | Overall N = 33,485[1] | Normal BP N = 18,699[1] | High-Normal N = 3,911[1] | IDH N = 2,317[1] | ISH N = 1,314[1] | SDH N = 1,781[1] | Diagnosed N = 5,463[1] | p-value[2] |
|---|---|---|---|---|---|---|---|---|
| **Social security (%)** | | | | | | | | <0.001 |
| Affiliated | 17,677 (53%) | 9,474 (51%) | 1,987 (51%) | 1,226 (53%) | 717 (55%) | 902 (51%) | 3,371 (62%) | |
| Not affiliated | 15,759 (47%) | 9,197 (49%) | 1,918 (49%) | 1,089 (47%) | 596 (45%) | 876 (49%) | 2,083 (38%) | |
| Missing | 49 | 28 | 6 | 2 | 1 | 3 | 9 | |
| **High DISLI (%)** | 8,787 (26%) | 5,101 (27%) | 1,080 (28%) | 601 (26%) | 367 (28%) | 476 (27%) | 1,162 (21%) | <0.001 |
| **SBP (mmHg)** | 120 (110, 130) | 112 (109, 120) | 130 (128, 132) | 130 (120, 130) | 145 (140, 152) | 150 (140, 160) | 130 (120, 141) | <0.001 |
| **DBP (mmHg)** | 80 (70, 84) | 72 (70, 80) | 81 (78, 86) | 90 (90, 94) | 80 (72, 83) | 97 (90, 100) | 80 (74, 90) | <0.001 |
| **WHtR** | 0.59 (0.54, 0.65) | 0.57 (0.52, 0.63) | 0.60 (0.55, 0.66) | 0.60 (0.55, 0.65) | 0.62 (0.56, 0.67) | 0.63 (0.58, 0.69) | 0.63 (0.58, 0.70) | <0.001 |
| Missing | 625 | 396 | 67 | 41 | 28 | 25 | 68 | |
| **BMI (kg/m2)** | 27.5 (24.4, 31.0) | 26.6 (23.7, 29.8) | 27.8 (25.0, 31.5) | 28.7 (25.8, 31.9) | 27.7 (24.6, 31.0) | 29.3 (26.2, 33.0) | 29.5 (26.2, 33.4) | <0.001 |
| **BMI-defined obesity (%)** | 10,364 (31%) | 4,445 (24%) | 1,311 (34%) | 891 (39%) | 410 (32%) | 799 (45%) | 2,508 (46%) | <0.001 |
| Missing | 288 | 135 | 31 | 17 | 22 | 20 | 63 | |
| **FPG (mg/dL)** | 94 (85, 107) | 91 (83, 101) | 95 (86, 109) | 96 (86, 107) | 100 (90, 135) | 100 (90, 118) | 99 (88, 122) | <0.001 |
| Missing | 23,585 | 13,235 | 2,761 | 1,590 | 943 | 1,255 | 3,801 | |
| **Prior diabetes diagnosis (%)** | 2,289 (6.8%) | 678 (3.6%) | 265 (6.8%) | 114 (4.9%) | 181 (14%) | 150 (8.4%) | 901 (16%) | <0.001 |
| **Prior CKD diagnosis (%)** | 356 (1.1%) | 146 (0.8%) | 30 (0.8%) | 20 (0.9%) | 18 (1.4%) | 17 (1.0%) | 125 (2.3%) | <0.001 |
| **Prior CVD diagnosis (%)** | 771 (2.3%) | 207 (1.1%) | 63 (1.6%) | 29 (1.3%) | 34 (2.6%) | 29 (1.6%) | 409 (7.5%) | <0.001 |

[1]Median (IQR); n (%). [2]Kruskal-Wallis rank sum test; Pearson's Chi-squared test

**Supplementary Table 4. ENSANUT 2012 population characteristics by hypertension status**. Abbreviations. BP: Blood Pressure; IDH: Isolated Diastolic Hypertension; ISH: Isolated Systolic Hypertension; SDH: Systolic-Diastolic Hypertension; DISLI: Density Independent Social Lag Index; SBP: Systolic Blood Pressure; DBP: Diastolic Blood Pressure; WHtR: Waist-to-height ratio; BMI: Body-mass index; FPG, Fasting Plasma Glucose; CKD, Chronic Kidney Disease; CVD, Cardiovascular Disease.

| ENSANUT 2012 Characteristic | Overall N = 10,832[1] | Normal BP N = 6,054[1] | High-Normal N = 1,168[1] | IDH N = 739[1] | ISH N = 386[1] | SDH N = 587[1] | Diagnosed N = 1,898[1] | p-value[2] |
|---|---|---|---|---|---|---|---|---|
| **Age (years)** | 42 (32, 57) | 36 (28, 47) | 46 (35, 60) | 42 (33, 52) | 62 (50, 71) | 52 (41, 65) | 57 (46, 68) | <0.001 |
| **Female (%)** | 6,575 (61%) | 3,897 (64%) | 571 (49%) | 356 (48%) | 173 (45%) | 276 (47%) | 1,302 (69%) | <0.001 |
| **Smoking (%)** | | | | | | | | <0.001 |
| Never | 5,969 (55%) | 3,474 (57%) | 567 (49%) | 349 (47%) | 190 (49%) | 275 (47%) | 1,114 (59%) | |
| Former | 2,966 (27%) | 1,475 (24%) | 358 (31%) | 221 (30%) | 136 (35%) | 188 (32%) | 588 (31%) | |
| Current | 1,886 (17%) | 1,101 (18%) | 243 (21%) | 168 (23%) | 60 (16%) | 123 (21%) | 191 (10%) | |
| Missing | 11 | 4 | 0 | 1 | 0 | 1 | 5 | |
| **Alcohol intake (%)** | | | | | | | | |
| Not currently | 5,761 (54%) | 3,067 (51%) | 592 (51%) | 348 (47%) | 223 (59%) | 308 (53%) | 1,223 (66%) | |
| Less than daily | 4,847 (45%) | 2,881 (48%) | 555 (48%) | 372 (51%) | 150 (39%) | 261 (45%) | 628 (34%) | |
| Daily | 116 (1.1%) | 54 (0.9%) | 12 (1.0%) | 15 (2.0%) | 8 (2.1%) | 15 (2.6%) | 12 (0.6%) | |
| Missing | 108 | 52 | 9 | 4 | 5 | 3 | 35 | |
| **Education level (%)** | | | | | | | | <0.001 |
| None | 1,262 (12%) | 530 (8.8%) | 155 (13%) | 70 (9.5%) | 98 (25%) | 93 (16%) | 316 (17%) | |
| Primary | 4,507 (42%) | 2,193 (36%) | 514 (44%) | 303 (41%) | 204 (53%) | 299 (51%) | 994 (52%) | |
| Secondary | 4,164 (38%) | 2,745 (45%) | 412 (35%) | 304 (41%) | 71 (18%) | 153 (26%) | 479 (25%) | |
| Tertiary | 899 (8.3%) | 586 (9.7%) | 87 (7.4%) | 62 (8.4%) | 13 (3.4%) | 42 (7.2%) | 109 (5.7%) | |
| **Indigenous identity (%)** | 1,171 (11%) | 710 (12%) | 130 (11%) | 59 (8.0%) | 61 (16%) | 65 (11%) | 146 (7.7%) | <0.001 |

| **ENSANUT 2012 Characteristic** | **Overall** N = 10,832[1] | **Normal BP** N = 6,054[1] | **High-Normal** N = 1,168[1] | **IDH** N = 739[1] | **ISH** N = 386[1] | **SDH** N = 587[1] | **Diagnosed** N = 1,898[1] | **p-value**[2] |
|---|---|---|---|---|---|---|---|---|
| **Social security (%)** | | | | | | | | <0.001 |
| Affiliated | 8,717 (81%) | 4,796 (79%) | 941 (81%) | 587 (79%) | 302 (78%) | 449 (76%) | 1,642 (87%) | |
| Not affiliated | 2,106 (19%) | 1,253 (21%) | 225 (19%) | 152 (21%) | 84 (22%) | 138 (24%) | 254 (13%) | |
| Missing | 9 | 5 | 2 | 0 | 0 | 0 | 2 | |
| **High DISLI (%)** | 2,883 (27%) | 1,662 (27%) | 318 (27%) | 177 (24%) | 128 (33%) | 145 (25%) | 453 (24%) | <0.001 |
| **SBP (mmHg)** | 120 (110, 130) | 114 (109, 120) | 130 (127, 132) | 130 (121, 130) | 145 (140, 155) | 150 (140, 160) | 130 (120, 147) | <0.001 |
| **DBP (mmHg)** | 80 (70, 85) | 74 (70, 80) | 82 (78, 86) | 90 (90, 95) | 80 (75, 84) | 98 (90, 101) | 81 (75, 90) | <0.001 |
| **WHtR** | 0.59 (0.54, 0.65) | 0.57 (0.52, 0.63) | 0.60 (0.55, 0.65) | 0.60 (0.55, 0.66) | 0.61 (0.56, 0.67) | 0.62 (0.57, 0.67) | 0.64 (0.58, 0.70) | <0.001 |
| Missing | 429 | 359 | 11 | 19 | 1 | 6 | 33 | |
| **BMI (kg/m2)** | 27.8 (24.5, 31.4) | 26.9 (23.7, 30.4) | 28.2 (25.2, 31.6) | 28.8 (25.6, 32.2) | 27.9 (24.7, 31.2) | 29.2 (25.8, 32.6) | 29.7 (26.2, 33.6) | <0.001 |
| **BMI-defined obesity (%)** | 3,635 (34%) | 1,633 (27%) | 411 (35%) | 307 (42%) | 135 (35%) | 253 (43%) | 896 (47%) | <0.001 |
| **FPG (mg/dL)** | 95 (87, 105) | 93 (86, 101) | 96 (88, 107) | 97 (88, 106) | 100 (91, 117) | 98 (91, 113) | 101 (91, 120) | <0.001 |
| Missing | 2,135 | 1,094 | 237 | 157 | 87 | 141 | 419 | |
| **Prior diabetes diagnosis (%)** | 1,039 (9.6%) | 256 (4.2%) | 102 (8.7%) | 57 (7.7%) | 62 (16%) | 56 (9.5%) | 506 (27%) | <0.001 |
| **Prior CKD diagnosis (%)** | *Data not available for this cycle* | | | | | | | |
| Missing | - | - | - | - | - | - | - | - |
| **Prior CVD diagnosis (%)** | 452 (4.2%) | 151 (2.5%) | 33 (2.8%) | 31 (4.2%) | 20 (5.2%) | 17 (2.9%) | 200 (11%) | <0.001 |

[1]Median (IQR); n (%). [2]Kruskal-Wallis rank sum test; Pearson's Chi-squared test

**Supplementary Table 5. ENSANUT 2016 population characteristics by hypertension status**. Abbreviations. BP: Blood Pressure; IDH: Isolated Diastolic Hypertension; ISH: Isolated Systolic Hypertension; SDH: Systolic-Diastolic Hypertension; DISLI: Density Independent Social Lag Index; SBP: Systolic Blood Pressure; DBP: Diastolic Blood Pressure; WHtR: Waist-to-height ratio; BMI: Body-mass index; FPG, Fasting Plasma Glucose; CKD, Chronic Kidney Disease; CVD, Cardiovascular Disease.

| ENSANUT 2016 Characteristic | Overall N = 8,161[1] | Normal BP N = 5,132[1] | High-Normal N = 830[1] | IDH N = 130[1] | ISH N = 516[1] | SDH N = 193[1] | Diagnosed N = 1,360[1] | p-value[2] |
|---|---|---|---|---|---|---|---|---|
| **Age (years)** | 44 (33, 58) | 38 (29, 50) | 48 (38, 59) | 43 (33, 50) | 63 (50, 72) | 48 (40, 59) | 59 (49, 69) | <0.001 |
| **Female (%)** | 5,355 (66%) | 3,501 (68%) | 426 (51%) | 79 (61%) | 272 (53%) | 98 (51%) | 979 (72%) | <0.001 |
| **Smoking (%)** | | | | | | | | <0.001 |
| Never | 4,195 (52%) | 2,629 (51%) | 423 (51%) | 65 (50%) | 258 (50%) | 96 (50%) | 724 (53%) | |
| Former | 2,929 (36%) | 1,795 (35%) | 298 (36%) | 48 (37%) | 200 (39%) | 66 (34%) | 522 (38%) | |
| Current | 1,007 (12%) | 687 (13%) | 105 (13%) | 17 (13%) | 57 (11%) | 31 (16%) | 110 (8.1%) | |
| Missing | 30 | 21 | 4 | 0 | 1 | 0 | 4 | |
| **Alcohol intake (%)** | | | | | | | | |
| Not currently | 3,072 (66%) | 1,840 (63%) | 309 (61%) | 49 (62%) | 202 (67%) | 63 (54%) | 609 (79%) | |
| Less than daily | 1,574 (34%) | 1,048 (36%) | 193 (38%) | 30 (38%) | 97 (32%) | 51 (44%) | 155 (20%) | |
| Daily | 33 (0.7%) | 15 (0.5%) | 5 (1.0%) | 0 (0%) | 3 (1.0%) | 2 (1.7%) | 8 (1.0%) | |
| Missing | 3,482 | 2,229 | 323 | 51 | 214 | 77 | 588 | |
| **Education level (%)** | | | | | | | | <0.001 |
| None | 961 (12%) | 421 (8.2%) | 104 (13%) | 13 (10%) | 142 (28%) | 20 (10%) | 261 (19%) | |
| Primary | 3,015 (37%) | 1,624 (32%) | 366 (44%) | 40 (31%) | 247 (48%) | 88 (46%) | 650 (48%) | |
| Secondary | 3,569 (44%) | 2,624 (51%) | 305 (37%) | 65 (50%) | 108 (21%) | 73 (38%) | 394 (29%) | |
| Tertiary | 616 (7.5%) | 463 (9.0%) | 55 (6.6%) | 12 (9.2%) | 19 (3.7%) | 12 (6.2%) | 55 (4.0%) | |
| **Indigenous identity (%)** | 945 (12%) | 617 (12%) | 107 (13%) | 9 (6.9%) | 82 (16%) | 23 (12%) | 107 (7.9%) | <0.001 |

| **ENSANUT 2016 Characteristic** | **Overall** N = 8,161[1] | **Normal BP** N = 5,132[1] | **High-Normal** N = 830[1] | **IDH** N = 130[1] | **ISH** N = 516[1] | **SDH** N = 193[1] | **Diagnosed** N = 1,360[1] | **p-value**[2] |
|---|---|---|---|---|---|---|---|---|
| **Social security (%)** | | | | | | | | 0.048 |
| Affiliated | 4,764 (88%) | 2,854 (88%) | 517 (88%) | 81 (93%) | 324 (85%) | 119 (88%) | 869 (90%) | |
| Not affiliated | 621 (12%) | 374 (12%) | 72 (12%) | 6 (6.9%) | 59 (15%) | 17 (13%) | 93 (9.7%) | |
| Missing | 2,776 | 1,904 | 241 | 43 | 133 | 57 | 398 | |
| **High DISLI (%)** | 2,805 (34%) | 1,771 (35%) | 307 (37%) | 40 (31%) | 206 (40%) | 61 (32%) | 420 (31%) | 0.003 |
| **SBP (mmHg)** | 118 (108, 131) | 111 (104, 119) | 133 (130, 136) | 132 (126, 136) | 148 (144, 158) | 152 (146, 162) | 133 (119, 151) | <0.001 |
| **DBP (mmHg)** | 72 (65, 80) | 69 (63, 75) | 81 (75, 86) | 92 (91, 95) | 79 (72, 84) | 97 (93, 101) | 77 (68, 85) | <0.001 |
| **WHtR** | 0.60 (0.55, 0.66) | 0.59 (0.53, 0.64) | 0.62 (0.56, 0.67) | 0.64 (0.60, 0.71) | 0.62 (0.57, 0.68) | 0.64 (0.58, 0.68) | 0.65 (0.60, 0.70) | <0.001 |
| Missing | 305 | 221 | 13 | 7 | 9 | 3 | 52 | |
| **BMI (kg/m2)** | 28.0 (24.8, 31.7) | 27.4 (24.3, 30.8) | 28.5 (25.7, 32.2) | 31.5 (28.0, 36.1) | 27.8 (24.6, 32.0) | 30.1 (26.9, 33.5) | 30.0 (26.5, 33.7) | <0.001 |
| **BMI-defined obesity (%)** | 2,869 (35%) | 1,539 (30%) | 310 (38%) | 76 (60%) | 175 (34%) | 97 (51%) | 672 (50%) | <0.001 |
| Missing | 44 | 14 | 6 | 4 | 1 | 2 | 17 | |
| **FPG (mg/dL)** | 95 (88, 105) | 93 (87, 101) | 97 (90, 108) | 102 (92, 113) | 99 (91, 120) | 101 (94, 111) | 103 (93, 131) | <0.001 |
| Missing | 4,349 | 2,709 | 438 | 76 | 287 | 107 | 732 | |
| **Prior diabetes diagnosis (%)** | 892 (11%) | 288 (5.6%) | 91 (11%) | 8 (6.2%) | 76 (15%) | 13 (6.7%) | 416 (31%) | <0.001 |
| Missing | 12 | 10 | 2 | 0 | 0 | 0 | 0 | |
| **Prior CKD diagnosis (%)** | 98 (1.2%) | 39 (0.8%) | 4 (0.5%) | 1 (0.8%) | 5 (1.0%) | 2 (1.0%) | 47 (3.5%) | |
| Missing | 13 | 10 | 2 | 0 | 0 | 0 | 1 | |
| **Prior CVD diagnosis (%)** | 242 (3.0%) | 88 (1.7%) | 12 (1.4%) | 3 (2.3%) | 9 (1.7%) | 4 (2.1%) | 126 (9.3%) | |
| Missing | 13 | 10 | 2 | 0 | 0 | 0 | 1 | |

[1]Median (IQR); n (%). [2]Kruskal-Wallis rank sum test; Pearson's Chi-squared test

**Supplementary Table 6. ENSANUT 2018 population characteristics by hypertension status**. Abbreviations. BP: Blood Pressure; IDH: Isolated Diastolic Hypertension; ISH: Isolated Systolic Hypertension; SDH: Systolic-Diastolic Hypertension; DISLI: Density Independent Social Lag Index; SBP: Systolic Blood Pressure; DBP: Diastolic Blood Pressure; WHtR: Waist-to-height ratio; BMI: Body-mass index; FPG, Fasting Plasma Glucose; CKD, Chronic Kidney Disease; CVD, Cardiovascular Disease.

| ENSANUT 2018 Characteristic | Overall N = 14,953[1] | Normal BP N = 8,006[1] | High-Normal N = 1,776[1] | IDH N = 371[1] | ISH N = 996[1] | SDH N = 547[1] | Diagnosed N = 3,257[1] | p-value[2] |
|---|---|---|---|---|---|---|---|---|
| **Age (years)** | 44 (33, 59) | 38 (29, 48) | 45 (34, 58) | 41 (32, 48) | 63 (52, 73) | 49 (40, 58) | 59 (47, 69) | <0.001 |
| **Female (%)** | 8,571 (57%) | 4,981 (62%) | 715 (40%) | 177 (48%) | 419 (42%) | 205 (37%) | 2,074 (64%) | <0.001 |
| **Smoking (%)** | | | | | | | | <0.001 |
| Never | 9,323 (62%) | 5,109 (64%) | 1,007 (57%) | 218 (59%) | 574 (58%) | 335 (61%) | 2,080 (64%) | |
| Former | 3,249 (22%) | 1,562 (20%) | 418 (24%) | 75 (20%) | 266 (27%) | 111 (20%) | 817 (25%) | |
| Current | 2,348 (16%) | 1,317 (16%) | 348 (20%) | 78 (21%) | 154 (15%) | 99 (18%) | 352 (11%) | |
| Missing | 33 | 18 | 3 | 0 | 2 | 2 | 8 | |
| **Alcohol intake (%)** | | | | | | | | |
| Not currently | 9,698 (65%) | 5,084 (64%) | 1,026 (58%) | 203 (55%) | 669 (67%) | 299 (55%) | 2,417 (74%) | |
| Less than daily | 5,063 (34%) | 2,845 (36%) | 719 (41%) | 162 (44%) | 307 (31%) | 236 (43%) | 794 (24%) | |
| Daily | 186 (1.2%) | 76 (0.9%) | 30 (1.7%) | 6 (1.6%) | 19 (1.9%) | 12 (2.2%) | 43 (1.3%) | |
| Missing | 6 | 1 | 1 | 0 | 1 | 0 | 3 | |
| **Education level (%)** | | | | | | | | <0.001 |
| None | 1,177 (7.9%) | 358 (4.5%) | 140 (7.9%) | 19 (5.1%) | 169 (17%) | 61 (11%) | 430 (13%) | |
| Primary | 4,678 (31%) | 1,945 (24%) | 542 (31%) | 88 (24%) | 476 (48%) | 205 (37%) | 1,422 (44%) | |
| Secondary | 7,060 (47%) | 4,391 (55%) | 842 (47%) | 207 (56%) | 272 (27%) | 233 (43%) | 1,115 (34%) | |
| Tertiary | 2,038 (14%) | 1,312 (16%) | 252 (14%) | 57 (15%) | 79 (7.9%) | 48 (8.8%) | 290 (8.9%) | |
| **Indigenous identity (%)** | 1,338 (8.9%) | 732 (9.1%) | 164 (9.2%) | 19 (5.1%) | 130 (13%) | 52 (9.5%) | 241 (7.4%) | <0.001 |

| **ENSANUT 2018 Characteristic** | **Overall** N = 14,953[1] | **Normal BP** N = 8,006[1] | **High-Normal** N = 1,776[1] | **IDH** N = 371[1] | **ISH** N = 996[1] | **SDH** N = 547[1] | **Diagnosed** N = 3,257[1] | **p-value**[2] |
|---|---|---|---|---|---|---|---|---|
| **Social security (%)** | | | | | | | | <0.001 |
| Affiliated | 12,868 (86%) | 6,850 (86%) | 1,494 (84%) | 305 (83%) | 843 (85%) | 452 (83%) | 2,924 (90%) | |
| Not affiliated | 2,024 (14%) | 1,117 (14%) | 277 (16%) | 63 (17%) | 150 (15%) | 94 (17%) | 323 (9.9%) | |
| Missing | 61 | 39 | 5 | 3 | 3 | 1 | 10 | |
| **High DISLI (%)** | 4,221 (28%) | 2,307 (29%) | 528 (30%) | 71 (19%) | 308 (31%) | 114 (21%) | 893 (27%) | <0.001 |
| **SBP (mmHg)** | 122 (111, 135) | 113 (105, 121) | 132 (130, 135) | 133 (128, 137) | 148 (143, 158) | 154 (146, 167) | 133 (120, 150) | <0.001 |
| **DBP (mmHg)** | 75 (68, 83) | 71 (65, 77) | 83 (76, 87) | 93 (91, 96) | 80 (74, 85) | 97 (93, 103) | 78 (70, 87) | <0.001 |
| **WHtR** | 0.60 (0.55, 0.66) | 0.58 (0.53, 0.64) | 0.60 (0.55, 0.65) | 0.62 (0.57, 0.67) | 0.61 (0.57, 0.66) | 0.63 (0.58, 0.69) | 0.64 (0.59, 0.70) | <0.001 |
| Missing | 839 | 499 | 68 | 19 | 46 | 15 | 192 | |
| **BMI (kg/m2)** | 28.3 (25.1, 31.9) | 27.4 (24.3, 30.8) | 28.6 (25.6, 32.1) | 30.3 (26.8, 33.8) | 27.8 (24.8, 31.3) | 30.3 (27.2, 33.4) | 29.9 (26.8, 33.9) | <0.001 |
| **BMI-defined obesity (%)** | 5,414 (37%) | 2,381 (30%) | 687 (39%) | 194 (53%) | 309 (32%) | 285 (53%) | 1,558 (50%) | <0.001 |
| Missing | 266 | 66 | 32 | 5 | 34 | 7 | 122 | |
| **FPG (mg/dL)** | 91 (83, 103) | 89 (82, 97) | 92 (84, 105) | 93 (85, 103) | 95 (87, 115) | 95 (86, 112) | 96 (87, 116) | <0.001 |
| Missing | 3,427 | 1,836 | 426 | 80 | 228 | 125 | 732 | |
| **Prior diabetes diagnosis (%)** | 1,805 (12%) | 459 (5.7%) | 187 (11%) | 21 (5.7%) | 202 (20%) | 45 (8.2%) | 891 (27%) | <0.001 |
| **Prior CKD diagnosis (%)** | 181 (1.2%) | 47 (0.6%) | 15 (0.8%) | 1 (0.3%) | 10 (1.0%) | 8 (1.5%) | 100 (3.1%) | |
| **Prior CVD diagnosis (%)** | 432 (2.9%) | 118 (1.5%) | 18 (1.0%) | 5 (1.3%) | 18 (1.8%) | 13 (2.4%) | 260 (8.0%) | <0.001 |

[1]Median (IQR); n (%). [2]Kruskal-Wallis rank sum test; Pearson's Chi-squared test

**Supplementary Table 7. ENSANUT 2020 population characteristics by hypertension status**. Abbreviations. BP: Blood Pressure; IDH: Isolated Diastolic Hypertension; ISH: Isolated Systolic Hypertension; SDH: Systolic-Diastolic Hypertension; DISLI: Density Independent Social Lag Index; SBP: Systolic Blood Pressure; DBP: Diastolic Blood Pressure; WHtR: Waist-to-height ratio; BMI: Body-mass index; FPG, Fasting Plasma Glucose; CKD, Chronic Kidney Disease; CVD, Cardiovascular Disease.

| ENSANUT 2020 Characteristic | Overall N = 9,921[1] | Normal BP N = 5,755[1] | High-Normal N = 1,151[1] | IDH N = 302[1] | ISH N = 662[1] | SDH N = 403[1] | Diagnosed N = 1,648[1] | p-value[2] |
|---|---|---|---|---|---|---|---|---|
| **Age (years)** | 44 (31, 58) | 37 (27, 48) | 46 (35, 58) | 41 (33, 48) | 62 (52, 70) | 49 (43, 59) | 61 (51, 70) | <0.001 |
| **Female (%)** | 5,919 (60%) | 3,664 (64%) | 523 (45%) | 143 (47%) | 318 (48%) | 184 (46%) | 1,087 (66%) | <0.001 |
| **Smoking (%)** | | | | | | | | <0.001 |
| Never | 4,304 (66%) | 2,516 (67%) | 469 (63%) | 132 (60%) | 287 (63%) | 173 (63%) | 727 (69%) | |
| Former | 1,202 (19%) | 622 (17%) | 137 (18%) | 50 (23%) | 99 (22%) | 57 (21%) | 237 (22%) | |
| Current | 975 (15%) | 599 (16%) | 135 (18%) | 37 (17%) | 68 (15%) | 43 (16%) | 93 (8.8%) | |
| Missing | 3,440 | 2,018 | 410 | 83 | 208 | 130 | 591 | |
| **Alcohol intake (%)** | | | | | | | | |
| Not currently | 3,131 (48%) | 1,643 (44%) | 347 (47%) | 90 (41%) | 253 (56%) | 140 (51%) | 658 (62%) | |
| Less than daily | 3,278 (51%) | 2,065 (55%) | 384 (52%) | 125 (57%) | 192 (42%) | 125 (46%) | 387 (37%) | |
| Daily | 73 (1.1%) | 30 (0.8%) | 9 (1.2%) | 4 (1.8%) | 10 (2.2%) | 8 (2.9%) | 12 (1.1%) | |
| Missing | 3,439 | 2,017 | 411 | 83 | 207 | 130 | 591 | |
| **Education level (%)** | | | | | | | | <0.001 |
| None | 607 (6.1%) | 200 (3.5%) | 81 (7.0%) | 7 (2.3%) | 105 (16%) | 38 (9.4%) | 176 (11%) | |
| Primary | 2,605 (26%) | 1,109 (19%) | 326 (28%) | 65 (22%) | 301 (45%) | 132 (33%) | 672 (41%) | |
| Secondary | 4,776 (48%) | 3,089 (54%) | 532 (46%) | 162 (54%) | 197 (30%) | 183 (45%) | 613 (37%) | |
| Tertiary | 1,933 (19%) | 1,357 (24%) | 212 (18%) | 68 (23%) | 59 (8.9%) | 50 (12%) | 187 (11%) | |
| **Indigenous identity (%)** | 564 (5.7%) | 334 (5.8%) | 65 (5.6%) | 19 (6.3%) | 45 (6.8%) | 31 (7.7%) | 70 (4.2%) | 0.046 |

| ENSANUT 2020 Characteristic | Overall N = 9,921[1] | Normal BP N = 5,755[1] | High-Normal N = 1,151[1] | IDH N = 302[1] | ISH N = 662[1] | SDH N = 403[1] | Diagnosed N = 1,648[1] | p-value[2] |
|---|---|---|---|---|---|---|---|---|
| **Social security (%)** | | | | | | | | <0.001 |
| Affiliated | 4,680 (48%) | 2,532 (44%) | 514 (45%) | 142 (48%) | 305 (47%) | 178 (45%) | 1,009 (61%) | |
| Not affiliated | 5,162 (52%) | 3,178 (56%) | 624 (55%) | 156 (52%) | 348 (53%) | 222 (56%) | 634 (39%) | |
| Missing | 79 | 45 | 13 | 4 | 9 | 3 | 5 | |
| **High DISLI (%)** | 2,911 (29%) | 1,661 (29%) | 356 (31%) | 88 (29%) | 233 (35%) | 138 (34%) | 435 (26%) | <0.001 |
| **SBP (mmHg)** | 120 (109, 134) | 112 (104, 120) | 132 (129, 136) | 132 (127, 136) | 148 (144, 157) | 154 (146, 167) | 134 (121, 149) | <0.001 |
| **DBP (mmHg)** | 75 (68, 82) | 71 (65, 76) | 84 (76, 87) | 93 (91, 95) | 80 (74, 86) | 97 (93, 102) | 78 (70, 87) | <0.001 |
| **WHtR** | *Data not available for this cycle* | | | | | | | |
| Missing | - | - | - | - | - | - | - | - |
| **BMI (kg/m2)** | 28.2 (25.0, 32.0) | 27.5 (24.2, 31.1) | 28.7 (25.9, 32.0) | 30.4 (27.5, 34.0) | 27.8 (24.8, 31.0) | 29.8 (26.9, 33.9) | 30.2 (26.8, 34.1) | <0.001 |
| **BMI-defined obesity (%)** | 3,697 (37%) | 1,805 (31%) | 467 (41%) | 159 (53%) | 216 (33%) | 196 (49%) | 854 (52%) | <0.001 |
| **FPG (mg/dL)** | 89 (82, 100) | 87 (80, 95) | 91 (83, 105) | 95 (88, 116) | 94 (86, 119) | 94 (85, 111) | 94 (86, 118) | <0.001 |
| Missing | 7,716 | 4,466 | 886 | 248 | 537 | 326 | 1,253 | |
| **Prior diabetes diagnosis (%)** | 1,244 (13%) | 353 (6.1%) | 122 (11%) | 39 (13%) | 120 (18%) | 48 (12%) | 562 (34%) | <0.001 |
| **Prior CKD diagnosis (%)** | *Data not available for this cycle* | | | | | | | |
| Missing | - | - | - | - | - | - | - | - |
| **Prior CVD diagnosis (%)** | 160 (1.6%) | 38 (0.7%) | 15 (1.3%) | 1 (0.3%) | 5 (0.8%) | 4 (1.0%) | 97 (5.9%) | |

[1]Median (IQR); n (%). [2]Kruskal-Wallis rank sum test; Pearson's Chi-squared test

**Supplementary Table 8. ENSANUT 2021 population characteristics by hypertension status**. Abbreviations. BP: Blood Pressure; IDH: Isolated Diastolic Hypertension; ISH: Isolated Systolic Hypertension; SDH: Systolic-Diastolic Hypertension; DISLI: Density Independent Social Lag Index; SBP: Systolic Blood Pressure; DBP: Diastolic Blood Pressure; WHtR: Waist-to-height ratio; BMI: Body-mass index; FPG, Fasting Plasma Glucose; CKD, Chronic Kidney Disease; CVD, Cardiovascular Disease.

| ENSANUT 2021 Characteristic | Overall N = 8,209[1] | Normal BP N = 4,972[1] | High-Normal N = 812[1] | IDH N = 170[1] | ISH N = 400[1] | SDH N = 226[1] | Diagnosed N = 1,629[1] | p-value[2] |
|---|---|---|---|---|---|---|---|---|
| **Age (years)** | 45 (33, 59) | 39 (29, 50) | 48 (34, 60) | 39 (32, 49) | 62 (51, 71) | 49 (42, 60) | 60 (49, 70) | <0.001 |
| **Female (%)** | 5,284 (64%) | 3,376 (68%) | 378 (47%) | 97 (57%) | 215 (54%) | 95 (42%) | 1,123 (69%) | <0.001 |
| **Smoking (%)** | | | | | | | | <0.001 |
| Never | 5,530 (68%) | 3,347 (67%) | 504 (62%) | 103 (61%) | 290 (73%) | 143 (64%) | 1,143 (70%) | |
| Former | 1,413 (17%) | 829 (17%) | 141 (17%) | 28 (17%) | 63 (16%) | 32 (14%) | 320 (20%) | |
| Current | 1,238 (15%) | 783 (16%) | 162 (20%) | 37 (22%) | 45 (11%) | 49 (22%) | 162 (10.0%) | |
| Missing | 28 | 13 | 5 | 2 | 2 | 2 | 4 | |
| **Alcohol intake (%)** | | | | | | | | |
| Not currently | 4,441 (54%) | 2,511 (51%) | 407 (50%) | 82 (49%) | 240 (60%) | 120 (53%) | 1,081 (66%) | |
| Less than daily | 3,684 (45%) | 2,421 (49%) | 389 (48%) | 86 (51%) | 153 (38%) | 102 (45%) | 533 (33%) | |
| Daily | 71 (0.9%) | 33 (0.7%) | 13 (1.6%) | 1 (0.6%) | 7 (1.8%) | 4 (1.8%) | 13 (0.8%) | |
| Missing | 13 | 7 | 3 | 1 | 0 | 0 | 2 | |
| **Education level (%)** | | | | | | | | <0.001 |
| None | 527 (6.4%) | 196 (3.9%) | 58 (7.1%) | 5 (2.9%) | 60 (15%) | 16 (7.1%) | 192 (12%) | |
| Primary | 2,322 (28%) | 1,084 (22%) | 251 (31%) | 43 (25%) | 182 (46%) | 81 (36%) | 681 (42%) | |
| Secondary | 4,078 (50%) | 2,760 (56%) | 375 (46%) | 98 (58%) | 128 (32%) | 101 (45%) | 616 (38%) | |
| Tertiary | 1,282 (16%) | 932 (19%) | 128 (16%) | 24 (14%) | 30 (7.5%) | 28 (12%) | 140 (8.6%) | |
| **Indigenous identity (%)** | 382 (4.7%) | 225 (4.5%) | 43 (5.3%) | 9 (5.3%) | 37 (9.3%) | 14 (6.2%) | 54 (3.3%) | <0.001 |

| **ENSANUT 2021 Characteristic** | **Overall** N = 8,209[1] | **Normal BP** N = 4,972[1] | **High-Normal** N = 812[1] | **IDH** N = 170[1] | **ISH** N = 400[1] | **SDH** N = 226[1] | **Diagnosed** N = 1,629[1] | **p-value**[2] |
|---|---|---|---|---|---|---|---|---|
| **Social security (%)** | | | | | | | | <0.001 |
| Affiliated | 3,619 (44%) | 2,075 (42%) | 360 (44%) | 61 (36%) | 170 (43%) | 90 (40%) | 863 (53%) | |
| Not affiliated | 4,570 (56%) | 2,883 (58%) | 449 (56%) | 109 (64%) | 228 (57%) | 136 (60%) | 765 (47%) | |
| Missing | 20 | 14 | 3 | 0 | 2 | 0 | 1 | |
| **High DISLI (%)** | 1,453 (18%) | 902 (18%) | 137 (17%) | 34 (20%) | 78 (20%) | 44 (19%) | 258 (16%) | 0.2 |
| **SBP (mmHg)** | 118 (108, 131) | 111 (103, 119) | 133 (130, 136) | 133 (128, 136) | 147 (143, 156) | 153 (146, 163) | 130 (118, 146) | <0.001 |
| **DBP (mmHg)** | 73 (66, 81) | 70 (64, 76) | 82 (75, 86) | 93 (91, 95) | 81 (75, 86) | 96 (93, 101) | 76 (68, 85) | <0.001 |
| **WHtR** | 0.62 (0.56, 0.68) | 0.59 (0.54, 0.66) | 0.62 (0.57, 0.67) | 0.65 (0.59, 0.70) | 0.62 (0.57, 0.70) | 0.64 (0.59, 0.70) | 0.67 (0.61, 0.73) | <0.001 |
| Missing | 353 | 264 | 10 | 2 | 13 | 2 | 62 | |
| **BMI (kg/m2)** | 28.7 (25.2, 32.6) | 27.8 (24.4, 31.6) | 29.2 (26.2, 32.8) | 31.7 (28.4, 35.0) | 28.2 (24.7, 32.2) | 30.4 (27.0, 34.3) | 30.8 (27.4, 35.1) | <0.001 |
| **BMI-defined obesity (%)** | 3,303 (41%) | 1,704 (34%) | 355 (44%) | 101 (60%) | 145 (37%) | 117 (52%) | 881 (56%) | <0.001 |
| Missing | 89 | 17 | 3 | 1 | 12 | 2 | 54 | |
| **FPG (mg/dL)** | 93 (86, 104) | 92 (85, 100) | 94 (88, 103) | 94 (86, 110) | 98 (89, 124) | 94 (88, 101) | 97 (88, 118) | <0.001 |
| Missing | 6,096 | 3,706 | 600 | 111 | 293 | 169 | 1,217 | |
| **Prior diabetes diagnosis (%)** | 1,086 (13%) | 344 (6.9%) | 95 (12%) | 11 (6.5%) | 80 (20%) | 24 (11%) | 532 (33%) | <0.001 |
| **Prior CKD diagnosis (%)** | 67 (0.8%) | 20 (0.4%) | 5 (0.6%) | 0 (0%) | 0 (0%) | 1 (0.4%) | 41 (2.5%) | |
| **Prior CVD diagnosis (%)** | 131 (1.6%) | 43 (0.9%) | 10 (1.2%) | 2 (1.2%) | 1 (0.3%) | 5 (2.2%) | 70 (4.3%) | |

[1]Median (IQR); n (%). [2]Kruskal-Wallis rank sum test; Pearson's Chi-squared test

**Supplementary Table 9. ENSANUT 2022 population characteristics by hypertension status**. Abbreviations. BP: Blood Pressure; IDH: Isolated Diastolic Hypertension; ISH: Isolated Systolic Hypertension; SDH: Systolic-Diastolic Hypertension; DISLI: Density Independent Social Lag Index; SBP: Systolic Blood Pressure; DBP: Diastolic Blood Pressure; WHtR: Waist-to-height ratio; BMI: Body-mass index; FPG, Fasting Plasma Glucose; CKD, Chronic Kidney Disease; CVD, Cardiovascular Disease.

| ENSANUT 2022 Characteristic | Overall N = 8,775[1] | Normal BP N = 4,997[1] | High-Normal N = 923[1] | IDH N = 162[1] | ISH N = 595[1] | SDH N = 302[1] | Diagnosed N = 1,796[1] | p-value[2] |
|---|---|---|---|---|---|---|---|---|
| **Age (years)** | 47 (33, 60) | 39 (29, 51) | 49 (39, 60) | 41 (32, 50) | 63 (52, 71) | 50 (41, 59) | 61 (50, 70) | <0.001 |
| **Female (%)** | 5,575 (64%) | 3,377 (68%) | 439 (48%) | 71 (44%) | 308 (52%) | 133 (44%) | 1,247 (69%) | <0.001 |
| **Smoking (%)** | | | | | | | | <0.001 |
| Never | 5,933 (68%) | 3,391 (68%) | 587 (64%) | 105 (65%) | 392 (66%) | 191 (63%) | 1,267 (71%) | |
| Former | 1,463 (17%) | 767 (15%) | 161 (17%) | 19 (12%) | 113 (19%) | 66 (22%) | 337 (19%) | |
| Current | 1,362 (16%) | 826 (17%) | 173 (19%) | 38 (23%) | 90 (15%) | 45 (15%) | 190 (11%) | |
| Missing | 17 | 13 | 2 | 0 | 0 | 0 | 2 | |
| **Alcohol intake (%)** | | | | | | | | |
| Not currently | 4,475 (51%) | 2,340 (47%) | 423 (46%) | 63 (39%) | 347 (58%) | 134 (44%) | 1,168 (65%) | |
| Less than daily | 4,151 (47%) | 2,589 (52%) | 477 (52%) | 93 (57%) | 233 (39%) | 154 (51%) | 605 (34%) | |
| Daily | 146 (1.7%) | 66 (1.3%) | 23 (2.5%) | 6 (3.7%) | 15 (2.5%) | 14 (4.6%) | 22 (1.2%) | |
| Missing | 3 | 2 | 0 | 0 | 0 | 0 | 1 | |
| **Education level (%)** | | | | | | | | <0.001 |
| None | 583 (6.6%) | 203 (4.1%) | 75 (8.1%) | 4 (2.5%) | 94 (16%) | 22 (7.3%) | 185 (10%) | |
| Primary | 2,520 (29%) | 1,056 (21%) | 276 (30%) | 33 (20%) | 282 (47%) | 95 (31%) | 778 (43%) | |
| Secondary | 4,347 (50%) | 2,785 (56%) | 445 (48%) | 92 (57%) | 178 (30%) | 150 (50%) | 697 (39%) | |
| Tertiary | 1,325 (15%) | 953 (19%) | 127 (14%) | 33 (20%) | 41 (6.9%) | 35 (12%) | 136 (7.6%) | |
| **Indigenous identity (%)** | 577 (6.6%) | 323 (6.5%) | 68 (7.4%) | 9 (5.6%) | 67 (11%) | 27 (8.9%) | 83 (4.6%) | <0.001 |

| **ENSANUT 2022 Characteristic** | **Overall** N = 8,775[1] | **Normal BP** N = 4,997[1] | **High-Normal** N = 923[1] | **IDH** N = 162[1] | **ISH** N = 595[1] | **SDH** N = 302[1] | **Diagnosed** N = 1,796[1] | **p-value**[2] |
|---|---|---|---|---|---|---|---|---|
| **Social security (%)** | | | | | | | | <0.001 |
| Affiliated | 4,163 (48%) | 2,224 (45%) | 430 (47%) | 85 (53%) | 263 (45%) | 130 (43%) | 1,031 (58%) | |
| Not affiliated | 4,542 (52%) | 2,734 (55%) | 480 (53%) | 76 (47%) | 326 (55%) | 171 (57%) | 755 (42%) | |
| Missing | 70 | 39 | 13 | 1 | 6 | 1 | 10 | |
| **High DISLI (%)** | 1,789 (20%) | 1,045 (21%) | 220 (24%) | 34 (21%) | 151 (25%) | 49 (16%) | 290 (16%) | <0.001 |
| **SBP (mmHg)** | 120 (109, 134) | 112 (104, 120) | 133 (130, 136) | 134 (128, 137) | 149 (143, 157) | 152 (145, 165) | 132 (119, 146) | <0.001 |
| **DBP (mmHg)** | 73 (66, 81) | 70 (64, 75) | 81 (75, 86) | 92 (91, 95) | 80 (73, 85) | 96 (93, 100) | 76 (68, 84) | <0.001 |
| **WHtR** | 0.62 (0.56, 0.68) | 0.60 (0.54, 0.66) | 0.62 (0.57, 0.69) | 0.64 (0.58, 0.69) | 0.63 (0.59, 0.69) | 0.65 (0.60, 0.71) | 0.67 (0.61, 0.73) | <0.001 |
| Missing | 406 | 279 | 20 | 3 | 22 | 7 | 75 | |
| **BMI (kg/m2)** | 28.6 (25.3, 32.5) | 27.8 (24.5, 31.4) | 29.5 (26.5, 32.8) | 30.8 (27.9, 33.8) | 28.4 (24.9, 31.8) | 30.3 (27.6, 34.2) | 30.3 (27.0, 34.5) | <0.001 |
| **BMI-defined obesity (%)** | 3,443 (40%) | 1,660 (33%) | 404 (44%) | 97 (60%) | 214 (37%) | 154 (52%) | 914 (53%) | <0.001 |
| Missing | 137 | 30 | 14 | 0 | 21 | 6 | 66 | |
| **FPG (mg/dL)** | 90 (81, 102) | 88 (78, 97) | 91 (82, 108) | 91 (85, 100) | 95 (84, 113) | 91 (81, 120) | 94 (84, 117) | <0.001 |
| Missing | 6,713 | 3,836 | 718 | 121 | 449 | 228 | 1,361 | |
| **Prior diabetes diagnosis (%)** | 1,220 (14%) | 377 (7.5%) | 104 (11%) | 8 (4.9%) | 102 (17%) | 25 (8.3%) | 604 (34%) | <0.001 |
| **Prior CKD diagnosis (%)** | 147 (1.7%) | 57 (1.1%) | 8 (0.9%) | 0 (0%) | 13 (2.2%) | 2 (0.7%) | 67 (3.7%) | |
| **Prior CVD diagnosis (%)** | 376 (4.3%) | 132 (2.6%) | 27 (2.9%) | 3 (1.9%) | 28 (4.7%) | 4 (1.3%) | 182 (10%) | <0.001 |

[1]Median (IQR); n (%). [2]Kruskal-Wallis rank sum test; Pearson's Chi-squared test

**Supplementary Table 10. ENSANUT 2023 population characteristics by hypertension status**. Abbreviations. BP: Blood Pressure; IDH: Isolated Diastolic Hypertension; ISH: Isolated Systolic Hypertension; SDH: Systolic-Diastolic Hypertension; DISLI: Density Independent Social Lag Index; SBP: Systolic Blood Pressure; DBP: Diastolic Blood Pressure; WHtR: Waist-to-height ratio; BMI: Body-mass index; FPG, Fasting Plasma Glucose; CKD, Chronic Kidney Disease; CVD, Cardiovascular Disease.

| ENSANUT 2023 Characteristic | Overall N = 3,151[1] | Normal BP N = 1,813[1] | High-Normal N = 292[1] | IDH N = 57[1] | ISH N = 181[1] | SDH N = 112[1] | Diagnosed N = 696[1] | p-value[2] |
|---|---|---|---|---|---|---|---|---|
| **Age (years)** | 47 (33, 61) | 39 (29, 51) | 50 (40, 63) | 41 (35, 48) | 64 (54, 71) | 52 (44, 60) | 61 (49, 69) | <0.001 |
| **Female (%)** | 1,982 (63%) | 1,236 (68%) | 143 (49%) | 27 (47%) | 77 (43%) | 40 (36%) | 459 (66%) | <0.001 |
| **Smoking (%)** | | | | | | | | <0.001 |
| Never | 2,106 (67%) | 1,239 (69%) | 177 (61%) | 36 (63%) | 100 (55%) | 71 (63%) | 483 (69%) | |
| Former | 539 (17%) | 272 (15%) | 61 (21%) | 9 (16%) | 47 (26%) | 21 (19%) | 129 (19%) | |
| Current | 496 (16%) | 294 (16%) | 53 (18%) | 12 (21%) | 34 (19%) | 20 (18%) | 83 (12%) | |
| Missing | 10 | 8 | 1 | 0 | 0 | 0 | 1 | |
| **Alcohol intake (%)** | | | | | | | | |
| Not currently | 1,520 (48%) | 793 (44%) | 137 (47%) | 24 (42%) | 94 (52%) | 51 (46%) | 421 (60%) | |
| Less than daily | 1,582 (50%) | 991 (55%) | 148 (51%) | 30 (53%) | 87 (48%) | 57 (51%) | 269 (39%) | |
| Daily | 46 (1.5%) | 26 (1.4%) | 7 (2.4%) | 3 (5.3%) | 0 (0%) | 4 (3.6%) | 6 (0.9%) | |
| Missing | 3 | 3 | 0 | 0 | 0 | 0 | 0 | |
| **Education level (%)** | | | | | | | | |
| None | 166 (5.3%) | 53 (2.9%) | 20 (6.8%) | 0 (0%) | 23 (13%) | 5 (4.5%) | 65 (9.3%) | |
| Primary | 807 (26%) | 342 (19%) | 88 (30%) | 19 (33%) | 73 (40%) | 35 (31%) | 250 (36%) | |
| Secondary | 1,658 (53%) | 1,066 (59%) | 141 (48%) | 30 (53%) | 67 (37%) | 61 (54%) | 293 (42%) | |
| Tertiary | 520 (17%) | 352 (19%) | 43 (15%) | 8 (14%) | 18 (9.9%) | 11 (9.8%) | 88 (13%) | |
| **Indigenous identity (%)** | 109 (3.5%) | 58 (3.2%) | 14 (4.8%) | 2 (3.5%) | 8 (4.4%) | 4 (3.6%) | 23 (3.3%) | 0.7 |

| ENSANUT 2023 Characteristic | Overall N = 3,151[1] | Normal BP N = 1,813[1] | High-Normal N = 292[1] | IDH N = 57[1] | ISH N = 181[1] | SDH N = 112[1] | Diagnosed N = 696[1] | p-value[2] |
|---|---|---|---|---|---|---|---|---|
| **Social security (%)** | | | | | | | | <0.001 |
| Affiliated | 1,499 (48%) | 815 (45%) | 135 (47%) | 20 (35%) | 87 (49%) | 52 (47%) | 390 (56%) | |
| Not affiliated | 1,621 (52%) | 977 (55%) | 152 (53%) | 37 (65%) | 92 (51%) | 58 (53%) | 305 (44%) | |
| Missing | 31 | 21 | 5 | 0 | 2 | 2 | 1 | |
| **High DISLI (%)** | 616 (20%) | 369 (20%) | 56 (19%) | 10 (18%) | 42 (23%) | 24 (21%) | 115 (17%) | 0.2 |
| **SBP (mmHg)** | 119 (108, 133) | 111 (104, 119) | 133 (130, 136) | 133 (126, 135) | 149 (143, 156) | 151 (145, 165) | 132 (118, 148) | <0.001 |
| **DBP (mmHg)** | 74 (66, 81) | 70 (64, 76) | 82 (75, 86) | 92 (91, 95) | 79 (73, 85) | 96 (93, 104) | 77 (68, 85) | <0.001 |
| **WHtR** | 0.62 (0.56, 0.69) | 0.60 (0.54, 0.66) | 0.63 (0.58, 0.70) | 0.63 (0.57, 0.70) | 0.62 (0.57, 0.70) | 0.63 (0.59, 0.69) | 0.67 (0.61, 0.73) | <0.001 |
| Missing | 166 | 122 | 6 | 2 | 4 | 2 | 30 | |
| **BMI (kg/m2)** | 28.8 (25.4, 32.5) | 28.0 (24.7, 31.6) | 29.2 (25.9, 33.1) | 29.8 (27.3, 33.9) | 27.7 (24.8, 31.3) | 30.5 (27.3, 32.9) | 30.5 (26.9, 34.8) | <0.001 |
| **BMI-defined obesity (%)** | 1,266 (41%) | 625 (35%) | 135 (46%) | 27 (48%) | 62 (35%) | 58 (53%) | 359 (53%) | <0.001 |
| Missing | 50 | 18 | 1 | 1 | 4 | 2 | 24 | |
| **FPG (mg/dL)** | 93 (86, 105) | 91 (85, 99) | 96 (86, 109) | 96 (84, 145) | 95 (85, 118) | 99 (91, 104) | 100 (91, 134) | <0.001 |
| Missing | 2,223 | 1,300 | 186 | 42 | 130 | 75 | 490 | |
| **Prior diabetes diagnosis (%)** | 425 (13%) | 125 (6.9%) | 28 (9.6%) | 5 (8.8%) | 35 (19%) | 15 (13%) | 217 (31%) | <0.001 |
| **Prior CKD diagnosis (%)** | 42 (1.3%) | 11 (0.6%) | 5 (1.7%) | 0 (0%) | 3 (1.7%) | 0 (0%) | 23 (3.3%) | <0.001 |
| **Prior CVD diagnosis (%)** | 141 (4.5%) | 54 (3.0%) | 9 (3.1%) | 4 (7.0%) | 7 (3.9%) | 1 (0.9%) | 66 (9.5%) | |

[1]Median (IQR); n (%). [2]Kruskal-Wallis rank sum test; Pearson's Chi-squared test

**Supplementary Table 11. ESC/ESH hypertension prevalence from 2000-2023.** Prevalence estimates of overall, diagnosed and undiagnosed hypertension, and high-normal blood pressure (BP) using definitions based on the European Society of Cardiology/European Society of Hypertension (ESC/ESH) guidelines. The table also shows trends over time (prevalence ratios [PR] per year) for these estimates from weighted Poisson regression models. All estimates were derived and weighted using ENSANUT's complex sampling designs from 2000-2023.

| ENSANUT cycle | Prevalence (95% confidence interval) | | | |
|---|---|---|---|---|
| | Overall hypertension | Undiagnosed Hypertension | Diagnosed Hypertension | High-normal BP |
| **2000** | 33.1 (32.2 - 34.0) | 20.7 (19.8 - 21.6) | 12.3 (11.7 - 12.9) | 10.8 (10.3 - 11.3) |
| **2006** | 31.8 (30.9 - 32.7) | 15.3 (14.6 - 16.0) | 16.6 (16.0 - 17.2) | 11.5 (10.9 - 12.1) |
| **2012** | 32.0 (30.7 - 33.3) | 15.2 (14.2 - 16.2) | 16.8 (15.7 - 17.9) | 10.3 (9.5 - 11.1) |
| **2016** | 25.2 (23.0 - 27.4) | 10.1 (8.7 - 11.5) | 15.1 (13.1 - 17.1) | 10.8 (9.1 - 12.5) |
| **2018** | 33.7 (32.4 - 35.0) | 12.1 (11.2 - 13.0) | 21.6 (20.4 - 22.8) | 12.0 (11.1 - 12.9) |
| **2020** | 28.8 (27.7 - 29.9) | 14.0 (13.1 - 14.9) | 14.7 (13.8 - 15.6) | 11.9 (11.1 - 12.8) |
| **2021** | 27.3 (25.8 - 28.8) | 9.9 (8.9 - 10.9) | 17.4 (16.3 - 18.5) | 10.6 (9.5 - 11.7) |
| **2022** | 28.2 (26.7 - 29.7) | 11.8 (10.7 - 12.9) | 16.4 (15.3 - 17.5) | 11.1 (10.1 - 12.1) |
| **2023** | 30.3 (27.5 - 33.1) | 11.0 (9.2 - 12.8) | 19.3 (16.8 - 21.8) | 9.3 (7.6 - 11.1) |
| **Trend per year PR (95% CI)** | 0.996 (0.995-0.998) | 0.970 (0.968-0.972) | 1.016 (1.014-1.017) | 0.993 (0.992-0.994) |

**Supplementary Table 12. ACC/AHA hypertension prevalence from 2000-2023.** Prevalence estimates of overall, diagnosed and undiagnosed hypertension, and elevated blood pressure (BP) using definitions based on the American College of Cardiology/American Heart Association (ACC/AHA) guidelines. The table also shows trends over time (prevalence ratios [PR] per year) for these estimates from weighted Poisson regression models. All estimates were derived and weighted using ENSANUT's complex sampling designs from 2000-2023.

| ENSANUT cycle | Prevalence (95% confidence interval) | | | |
|---|---|---|---|---|
| | Overall hypertension | Undiagnosed Hypertension | Diagnosed Hypertension | Elevated BP |
| **2000** | 68.3 (67.3 - 69.3) | 56.0 (55.1 - 56.9) | 12.3 (11.7 - 12.9) | 6.9 (6.5 - 7.3) |
| **2006** | 60.0 (59.0 - 61.0) | 43.4 (42.5 - 44.3) | 16.6 (16.0 - 17.2) | 9.9 (9.3 - 10.5) |
| **2012** | 60.1 (58.6 - 61.6) | 43.4 (41.9 - 44.9) | 16.8 (15.7 - 17.9) | 10.0 (9.1 - 10.9) |
| **2016** | 40.8 (38.1 - 43.5) | 25.7 (23.5 - 27.9) | 15.1 (13.1 - 17.1) | 10.7 (9.5 - 11.9) |
| **2018** | 52.1 (50.7 - 53.5) | 30.5 (29.3 - 31.7) | 21.6 (20.4 - 22.8) | 11.4 (10.6 - 12.2) |
| **2020** | 47.9 (46.6 - 49.2) | 33.2 (32.0 - 34.4) | 14.7 (13.8 - 15.6) | 10.8 (10.0 - 11.6) |
| **2021** | 44.4 (42.7 - 46.1) | 27.0 (25.5 - 28.5) | 17.4 (16.3 - 18.5) | 10.6 (9.7 - 11.5) |
| **2022** | 45.7 (44.1 - 47.3) | 29.3 (27.9 - 30.7) | 16.4 (15.3 - 17.5) | 10.6 (9.5 - 11.7) |
| **2023** | 41.0 (37.9 - 44.1) | 21.7 (18.9 - 24.4) | 19.3 (16.8 - 21.8) | 10.0 (7.4 - 12.6) |
| **Trend per year PR (95% CI)** | 1.018 (1.016-1.020) | 0.968 (0.967-0.969) | 1.016 (1.014-1.017) | 0.981 (0.980-0.982) |

**Supplementary Table 13. ESC/ESH hypertension prevalence by sex.** Weighted prevalence estimates of overall, diagnosed and undiagnosed hypertension, and high-normal BP stratified by sex using ESC/ESH definitions from 2000 to 2023.

| ENSANUT | Sex | Prevalence (95% confidence interval) | | | |
|---|---|---|---|---|---|
| | | Overall hypertension | Undiagnosed hypertension | Diagnosed hypertension | High-normal BP |
| 2000 | Men | 31.0 (30.1 - 31.9) | 15.5 (14.7 - 16.3) | 15.5 (14.8 - 16.2) | 8.7 (8.1 - 9.3) |
| | Women | 35.4 (33.7 - 37.1) | 26.6 (25.1 - 28.1) | 8.8 (8.0 - 9.6) | 13.0 (12.3 - 13.7) |
| 2006 | Men | 32.6 (31.2 - 34.0) | 20.0 (18.9 - 21.1) | 12.7 (11.8 - 13.6) | 14.9 (14.0 - 15.8) |
| | Women | 31.3 (30.3 - 32.3) | 12.0 (11.3 - 12.7) | 19.3 (18.4 - 20.2) | 9.2 (8.6 - 9.8) |
| 2012 | Men | 33.3 (31.3 - 35.3) | 19.1 (17.5 - 20.7) | 14.2 (12.6 - 15.8) | 12.5 (11.1 - 13.9) |
| | Women | 31.0 (29.2 - 32.8) | 12.2 (10.9 - 13.5) | 18.8 (17.4 - 20.2) | 8.6 (7.6 - 9.6) |
| 2016 | Men | 24.7 (21.3 - 28.1) | 12.7 (10.7 - 14.7) | 11.9 (8.9 - 14.9) | 15.5 (12.5 - 18.5) |
| | Women | 25.7 (23.1 - 28.3) | 7.6 (6.3 - 8.9) | 18.1 (15.6 - 20.6) | 6.3 (5.2 - 7.4) |
| 2018 | Men | 34.1 (32.2 - 36.0) | 16.2 (14.8 - 17.6) | 17.9 (16.2 - 19.6) | 16.1 (14.6 - 17.6) |
| | Women | 33.4 (31.6 - 35.2) | 9.1 (8.1 - 10.1) | 24.3 (22.7 - 25.9) | 9.0 (7.9 - 10.1) |
| 2020 | Men | 30.4 (28.6 - 32.1) | 18.0 (16.5 - 19.5) | 12.4 (11.1 - 13.7) | 16.0 (14.5 - 17.5) |
| | Women | 27.3 (25.9 - 28.7) | 10.4 (9.5 - 11.3) | 16.9 (15.7 - 18.0) | 8.2 (7.3 - 9.1) |
| 2021 | Men | 29.2 (26.9 - 31.5) | 12.9 (11.1 - 14.7) | 16.2 (14.5 - 17.9) | 15.0 (13.0 - 17.0) |
| | Women | 25.8 (24.1 - 27.5) | 7.5 (6.5 - 8.5) | 18.3 (16.9 - 19.7) | 7.0 (5.9 - 8.1) |
| 2022 | Men | 30.1 (28.0 - 32.2) | 15.2 (13.4 - 17.0) | 14.8 (13.1 - 16.5) | 15.3 (13.5 - 17.1) |
| | Women | 26.6 (24.7 - 28.5) | 8.8 (7.4 - 10.2) | 17.8 (16.4 - 19.2) | 7.5 (6.4 - 8.6) |
| 2023 | Men | 36.0 (32.1 - 39.9) | 16.9 (13.1 - 20.7) | 19.1 (15.5 - 22.7) | 11.6 (8.9 - 14.3) |
| | Women | 25.3 (21.9 - 28.7) | 5.8 (4.2 - 7.4) | 19.5 (16.8 - 22.2) | 7.3 (5.5 - 9.1) |

<u>**Supplementary Table 14**</u>**. ESC/ESH hypertension prevalence by age group.** Weighted prevalence estimates of overall, diagnosed and undiagnosed hypertension, and high-normal BP stratified by age group using ESC/ESH definitions from 2000 to 2023.

| ENSANUT | Age (years) | Prevalence (95% confidence interval) | | | |
|---|---|---|---|---|---|
| | | Overall hypertension | Undiagnosed hypertension | Diagnosed hypertension | High-normal BP |
| 2000 | 20-39 | 21.9 (20.8 - 23.0) | 15.8 (14.8 - 16.9) | 6.1 (5.5 - 6.7) | 9.4 (8.7 - 10.1) |
| | 40-59 | 45.5 (43.9 - 47.1) | 27.3 (25.9 - 28.7) | 18.3 (17.3 - 19.3) | 12.3 (11.2 - 13.4) |
| | ≥60 | 60.1 (58.0 - 62.2) | 30.5 (28.8 - 32.2) | 29.6 (27.7 - 31.5) | 13.9 (12.2 - 15.6) |
| 2006 | 20-39 | 18.0 (17.1 - 18.9) | 9.9 (9.2 - 10.6) | 8.1 (7.4 - 8.8) | 10.0 (9.3 - 10.7) |
| | 40-59 | 37.4 (35.8 - 39.0) | 18.1 (16.9 - 19.3) | 19.3 (18.2 - 20.4) | 12.9 (12.0 - 13.8) |
| | ≥60 | 59.6 (57.8 - 61.4) | 24.7 (23.0 - 26.4) | 35.0 (33.0 - 37.0) | 13.4 (12.1 - 14.7) |
| 2012 | 20-39 | 15.9 (14.3 - 17.5) | 10.4 (9.0 - 11.8) | 5.6 (4.7 - 6.5) | 8.3 (7.3 - 9.3) |
| | 40-59 | 39.3 (37.0 - 41.6) | 18.5 (16.6 - 20.4) | 20.8 (18.9 - 22.7) | 12.1 (10.4 - 13.8) |
| | ≥60 | 59.6 (56.4 - 62.8) | 21.5 (19.1 - 23.9) | 38.1 (35.1 - 41.1) | 12.4 (10.1 - 14.7) |
| 2016 | 20-39 | 10.5 (7.7 - 13.3) | 5.4 (3.9 - 6.9) | 5.1 (2.5 - 7.7) | 10.1 (7.2 - 13.0) |
| | 40-59 | 30.6 (27.6 - 33.6) | 11.7 (9.6 - 13.8) | 18.9 (16.3 - 21.5) | 12.6 (10.4 - 14.8) |
| | ≥60 | 58.6 (54.5 - 62.7) | 21.0 (16.2 - 25.8) | 37.6 (33.0 - 42.2) | 9.4 (7.5 - 11.3) |
| 2018 | 20-39 | 13.8 (12.4 - 15.2) | 6.9 (5.7 - 8.1) | 6.8 (5.8 - 7.8) | 10.5 (9.3 - 11.7) |
| | 40-59 | 36.6 (34.4 - 38.8) | 13.6 (12.0 - 15.2) | 22.9 (20.9 - 24.9) | 13.7 (12.3 - 15.1) |
| | ≥60 | 63.3 (60.4 - 66.2) | 18.5 (16.4 - 20.6) | 44.8 (41.7 - 47.9) | 11.7 (9.3 - 14.0) |
| 2020 | 20-39 | 10.1 (8.8 - 11.3) | 7.3 (6.2 - 8.4) | 2.8 (2.2 - 3.4) | 10.5 (9.2 - 11.8) |
| | 40-59 | 33.5 (31.7 - 35.3) | 17.3 (15.8 - 18.8) | 16.2 (14.7 - 17.7) | 13.9 (12.3 - 15.5) |
| | ≥60 | 62.8 (60.3 - 65.3) | 23.4 (21.2 - 25.6) | 39.4 (36.9 - 41.9) | 11.3 (9.8 - 12.8) |

| ENSANUT | Age (years) | Prevalence (95% confidence interval) | | | |
|---|---|---|---|---|---|
| | | **Overall hypertension** | **Undiagnosed hypertension** | **Diagnosed hypertension** | **High-normal BP** |
| **2021** | 20-39 | 10.8 (9.1 - 12.5) | 6.1 (4.7 - 7.5) | 4.7 (3.7 - 5.7) | 9.3 (7.8 - 10.9) |
| | 40-59 | 30.8 (28.7 - 33.0) | 11.3 (9.6 - 13.0) | 19.4 (17.6 - 21.2) | 11.8 (10.3 - 13.3) |
| | ≥60 | 57.7 (54.5 - 60.9) | 15.8 (13.4 - 18.2) | 42.0 (39.0 - 45.0) | 11.4 (9.1 - 13.7) |
| **2022** | 20-39 | 11.0 (9.4 - 12.6) | 6.3 (5.0 - 7.6) | 4.8 (3.8 - 5.8) | 9.4 (7.8 - 11.0) |
| | 40-59 | 30.8 (28.5 - 33.1) | 13.7 (11.9 - 15.5) | 17.1 (15.4 - 18.9) | 13.9 (12.2 - 15.6) |
| | ≥60 | 61.0 (58.3 - 63.7) | 20.6 (18.1 - 23.1) | 40.4 (37.4 - 43.4) | 10.4 (8.6 - 12.2) |
| **2023** | 20-39 | 11.4 (8.4 - 14.4) | 6.1 (4.0 - 8.2) | 5.3 (3.3 - 7.3) | 7.1 (4.8 - 9.4) |
| | 40-59 | 35.7 (30.9 - 40.5) | 12.2 (9.2 - 15.2) | 23.5 (19.0 - 28.0) | 10.7 (7.4 - 14.0) |
| | ≥60 | 60.7 (55.0 - 66.4) | 19.2 (15.1 - 23.3) | 41.6 (35.0 - 48.2) | 11.6 (7.7 - 15.5) |

**Supplementary Table 15. ESC/ESH hypertension prevalence by obesity status.** Weighted prevalence estimates of overall, diagnosed and undiagnosed hypertension, and high-normal BP stratified by obesity status using ESC/ESH definitions from 2000-2023.

| ENSANUT | Obesity status | Prevalence (95% confidence interval) | | | |
|---|---|---|---|---|---|
| | | Overall hypertension | Undiagnosed hypertension | Diagnosed hypertension | High-normal BP |
| 2000 | No Obesity | 27.7 (26.7 - 28.7) | 18.5 (17.6 - 19.4) | 9.2 (8.7 - 9.7) | 10.3 (9.8 - 10.8) |
| | Obesity | 49.5 (47.8 - 51.2) | 27.6 (26.1 - 29.1) | 21.8 (20.4 - 23.2) | 12.0 (10.9 - 13.1) |
| 2006 | No Obesity | 26.3 (25.2 - 27.4) | 13.4 (12.7 - 14.1) | 12.9 (12.1 - 13.7) | 10.9 (10.3 - 11.5) |
| | Obesity | 44.6 (43.1 - 46.1) | 19.7 (18.5 - 20.9) | 25.0 (23.7 - 26.3) | 12.9 (11.9 - 13.9) |
| 2012 | No Obesity | 26.7 (25.1 - 28.3) | 13.1 (12.0 - 14.2) | 13.6 (12.4 - 14.8) | 9.5 (8.6 - 10.4) |
| | Obesity | 42.8 (40.3 - 45.3) | 19.5 (17.5 - 21.5) | 23.3 (21.2 - 25.4) | 11.9 (10.3 - 13.5) |
| 2016 | No Obesity | 20.7 (18.2 - 23.2) | 8.9 (7.4 - 10.4) | 11.8 (9.5 - 14.1) | 9.1 (7.5 - 10.7) |
| | Obesity | 33.7 (29.4 - 38.0) | 12.4 (9.9 - 14.9) | 21.3 (18.1 - 24.6) | 14.1 (10.2 - 18.0) |
| 2018 | No Obesity | 27.8 (26.2 - 29.4) | 10.9 (9.9 - 11.9) | 16.9 (15.5 - 18.3) | 11.3 (10.1 - 12.5) |
| | Obesity | 42.6 (40.3 - 44.9) | 13.9 (12.2 - 15.6) | 28.6 (26.6 - 30.7) | 13.1 (11.6 - 14.6) |
| 2020 | No Obesity | 23.8 (22.5 - 25.1) | 12.9 (11.8 - 14.0) | 11.0 (10.1 - 11.9) | 11.2 (10.1 - 12.3) |
| | Obesity | 37.4 (35.3 - 39.5) | 16.1 (14.5 - 17.7) | 21.4 (19.6 - 23.1) | 13.1 (11.7 - 14.5) |
| 2021 | No Obesity | 21.3 (19.5 - 23.1) | 8.4 (7.2 - 9.6) | 12.9 (11.7 - 14.1) | 9.8 (8.6 - 11.1) |
| | Obesity | 35.9 (33.6 - 38.2) | 12.2 (10.4 - 14.0) | 23.7 (21.7 - 25.7) | 12.0 (10.5 - 13.5) |
| 2022 | No Obesity | 22.7 (20.9 - 24.4) | 10.4 (9.1 - 11.7) | 12.3 (11.1 - 13.6) | 10.0 (8.8 - 11.2) |
| | Obesity | 36.0 (32.7 - 39.3) | 13.8 (11.7 - 15.9) | 22.3 (20.1 - 24.5) | 13.2 (11.2 - 15.1) |
| 2023 | No Obesity | 24.6 (21.7 - 27.5) | 9.2 (6.9 - 11.5) | 15.4 (12.6 - 18.2) | 7.8 (6.0 - 9.6) |
| | Obesity | 37.4 (33.1 - 41.6) | 13.8 (10.9 - 16.7) | 23.6 (19.6 - 27.6) | 11.8 (8.7 - 14.9) |

**Supplementary Table 16. ESC/ESH hypertension prevalence by diabetes status.** Weighted prevalence estimates of overall, diagnosed and undiagnosed hypertension, and high-normal BP stratified by diabetes status using ESC/ESH definitions (2000-2023).

| ENSANUT | Diabetes status | Prevalence (95% confidence interval) | | | |
|---|---|---|---|---|---|
| | | Overall hypertension | Undiagnosed hypertension | Diagnosed hypertension | High-normal BP |
| 2000 | No Diabetes | 31.3 (30.3 - 32.3) | 20.4 (19.5 - 21.3) | 11.0 (10.4 - 11.6) | 10.7 (10.2 - 11.2) |
| | Diabetes | 59.8 (57.3 - 62.3) | 25.7 (23.5 - 27.9) | 34.1 (31.7 - 36.5) | 12.1 (10.2 - 14.0) |
| 2006 | No Diabetes | 30.1 (28.6 - 31.6) | 15.1 (13.9 - 16.3) | 14.9 (13.7 - 16.1) | 11.1 (10.1 - 12.1) |
| | Diabetes | 56.2 (53.2 - 59.2) | 18.7 (16.3 - 21.1) | 37.5 (34.4 - 40.6) | 12.9 (10.7 - 15.1) |
| 2012 | No Diabetes | 26.2 (24.7 - 27.7) | 14.0 (12.9 - 15.1) | 12.2 (11.1 - 13.2) | 10.3 (9.3 - 11.3) |
| | Diabetes | 62.2 (58.3 - 66.1) | 19.1 (15.9 - 22.3) | 43.1 (38.9 - 47.3) | 10.4 (8.2 - 12.6) |
| 2016 | No Diabetes | 18.9 (16.5 - 21.3) | 8.6 (6.7 - 10.5) | 10.3 (8.5 - 12.1) | 12.0 (9.3 - 14.7) |
| | Diabetes | 55.9 (50.9 - 60.9) | 12.9 (9.5 - 16.3) | 43.0 (37.2 - 48.8) | 9.3 (6.9 - 11.7) |
| 2018 | No Diabetes | 28.9 (27.2 - 30.6) | 11.6 (10.4 - 12.8) | 17.4 (16.0 - 18.8) | 11.7 (10.5 - 12.9) |
| | Diabetes | 58.7 (55.5 - 61.9) | 16.2 (13.7 - 18.7) | 42.5 (39.0 - 46.0) | 10.3 (8.4 - 12.2) |
| 2020 | No Diabetes | 22.6 (20.2 - 25.0) | 10.1 (8.4 - 11.8) | 12.6 (10.9 - 14.3) | 12.5 (10.6 - 14.4) |
| | Diabetes | 52.9 (47.0 - 58.8) | 19.2 (14.3 - 24.1) | 33.7 (27.9 - 39.5) | 13.8 (9.8 - 17.8) |
| 2021 | No Diabetes | 22.8 (20.4 - 25.2) | 10.0 (8.2 - 11.8) | 12.8 (10.8 - 14.8) | 10.6 (8.6 - 12.6) |
| | Diabetes | 58.1 (54.1 - 62.1) | 12.9 (9.6 - 16.2) | 45.2 (41.3 - 49.1) | 8.2 (6.1 - 10.3) |
| 2022 | No Diabetes | 24.1 (21.1 - 27.1) | 11.9 (9.5 - 14.3) | 12.3 (10.4 - 14.2) | 10.4 (8.0 - 12.8) |
| | Diabetes | 55.0 (51.2 - 58.8) | 12.8 (10.3 - 15.3) | 42.1 (38.3 - 45.9) | 9.8 (7.6 - 12.0) |
| 2023 | No Diabetes | 22.6 (18.1 - 27.1) | 8.5 (5.4 - 11.6) | 14.1 (10.8 - 17.4) | 11.3 (8.0 - 14.6) |
| | Diabetes | 68.4 (61.7 - 75.1) | 16.9 (10.6 - 23.2) | 51.4 (44.3 - 58.5) | 8.7 (4.6 - 12.8) |

#### SUPPLEMENTARY FIGURES

**Supplementary Figure 1. Flowchart of participant selection.** Flow diagram showing the selection of participants from ENSANUT 2000-2023 for each analysis, including inclusion criteria and the use of complex survey designs to expand results to a nationally representative population. For **Analysis 1** (hypertension prevalence), we included a total of 141,668 Mexican adults aged ≥20 years. For **Analysis 2**, we analyzed 36,158 adults with hypertension (both diagnosed and undiagnosed) to identify predictors of lack of diagnosis†, and 16,411 adults with previously diagnosed hypertension to identify predictors of lack of treatment‡. Information on hypertension treatment was not collected in 2020.

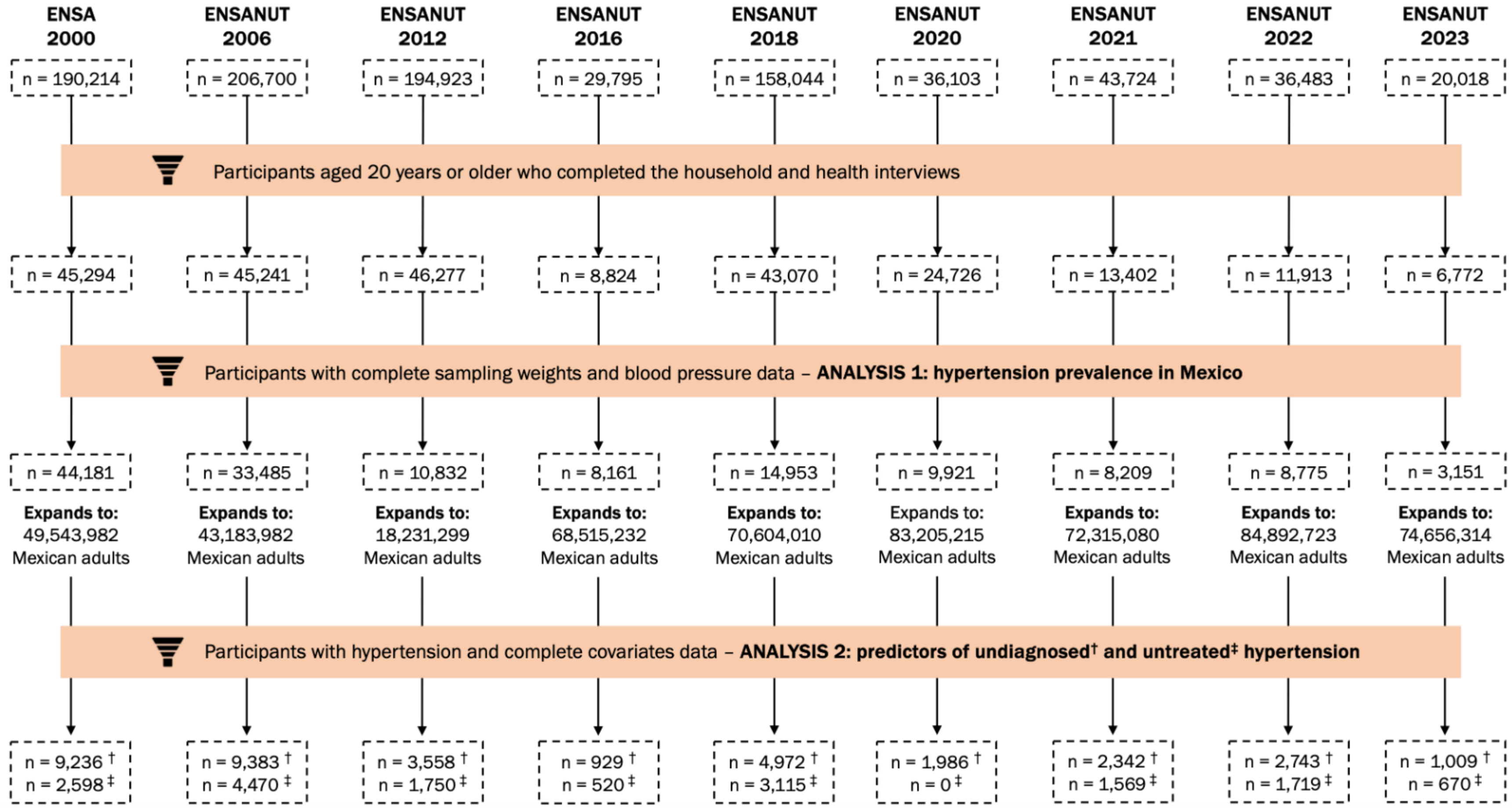

**Supplementary Figure 2. ACC/AHA hypertension prevalence trends**. Trends in overall, diagnosed and undiagnosed hypertension, and elevated blood pressure (BP) using data from ENSANUT from 2000-2023 in Mexico, based on ACC/AHA definitions.

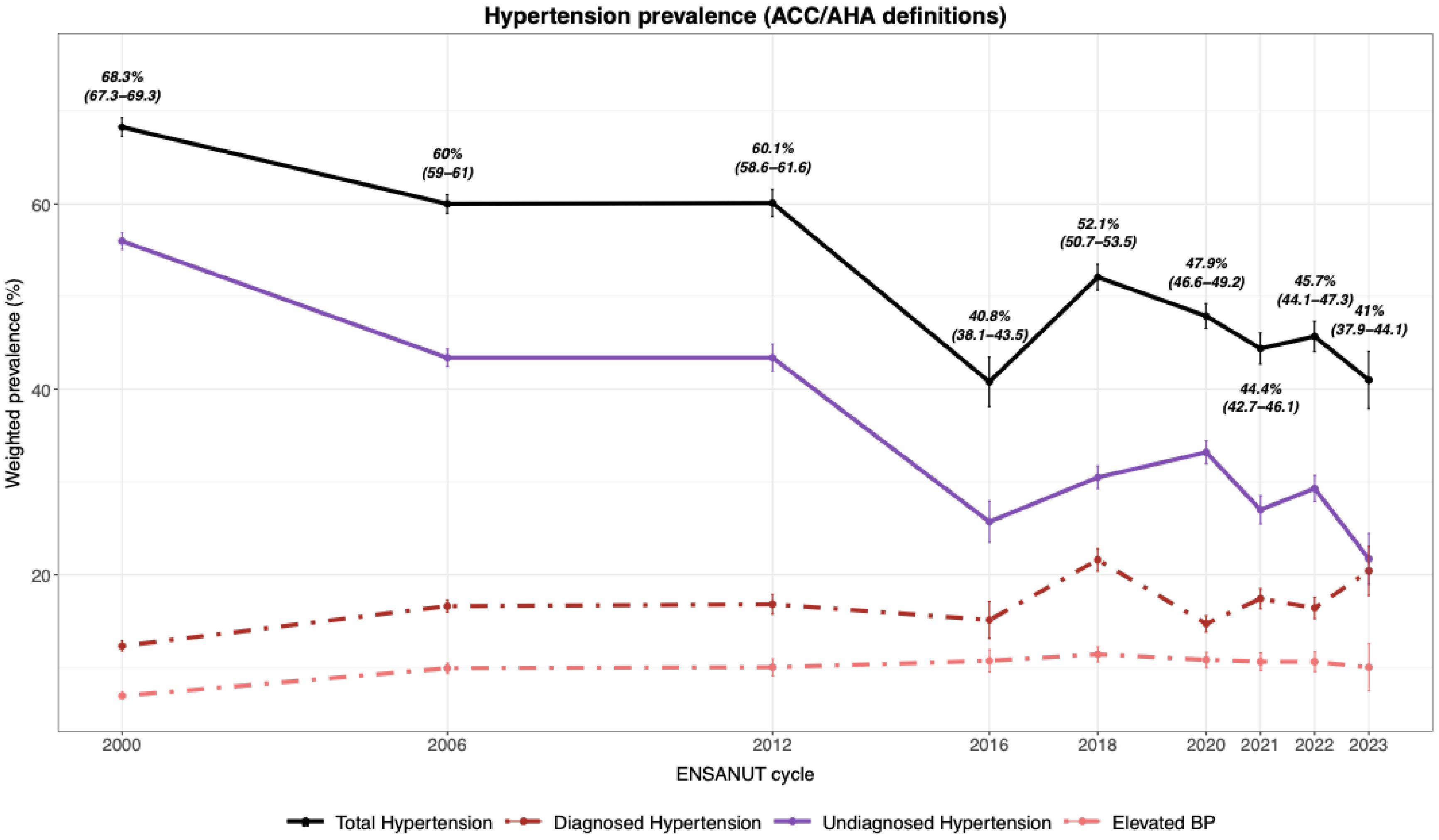

<u>**Supplementary Figure 3**</u>**. Prevalence of undiagnosed hypertension phenotypes (ACC/AHA)**. Prevalence of isolated systolic (ISH), isolated diastolic (IDH) and systolic-diastolic hypertension (SDH) in Mexican population using the ACC/AHA definitions.

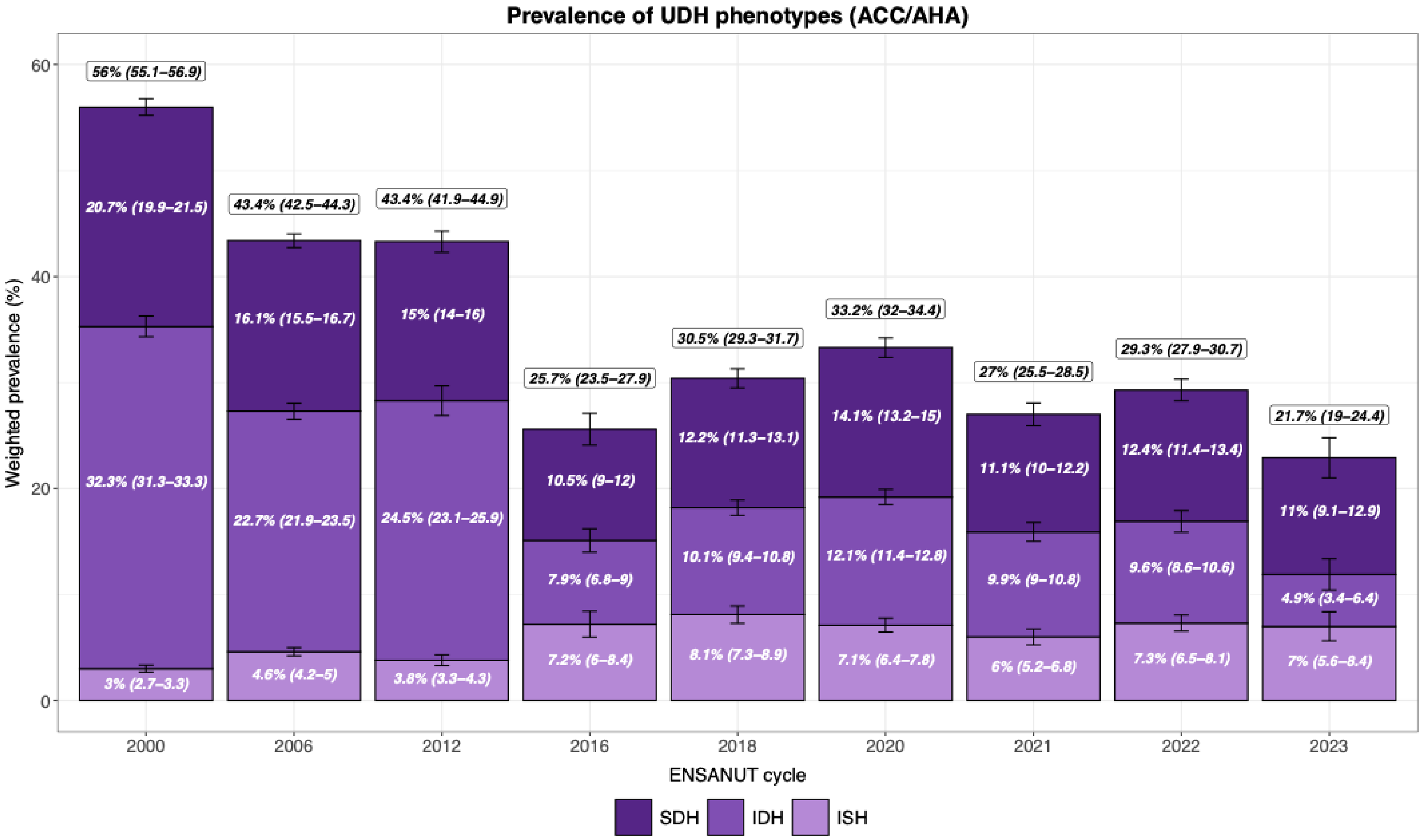

**Supplementary Figure 4. Percentage of phenotypes among UDH (ESC/ESH).** Estimated percentage of isolated systolic (ISH), isolated diastolic (IDH) and systolic-diastolic hypertension (SDH) within participants with undiagnosed hypertension (UDH) using the ESC/ESH definitions.

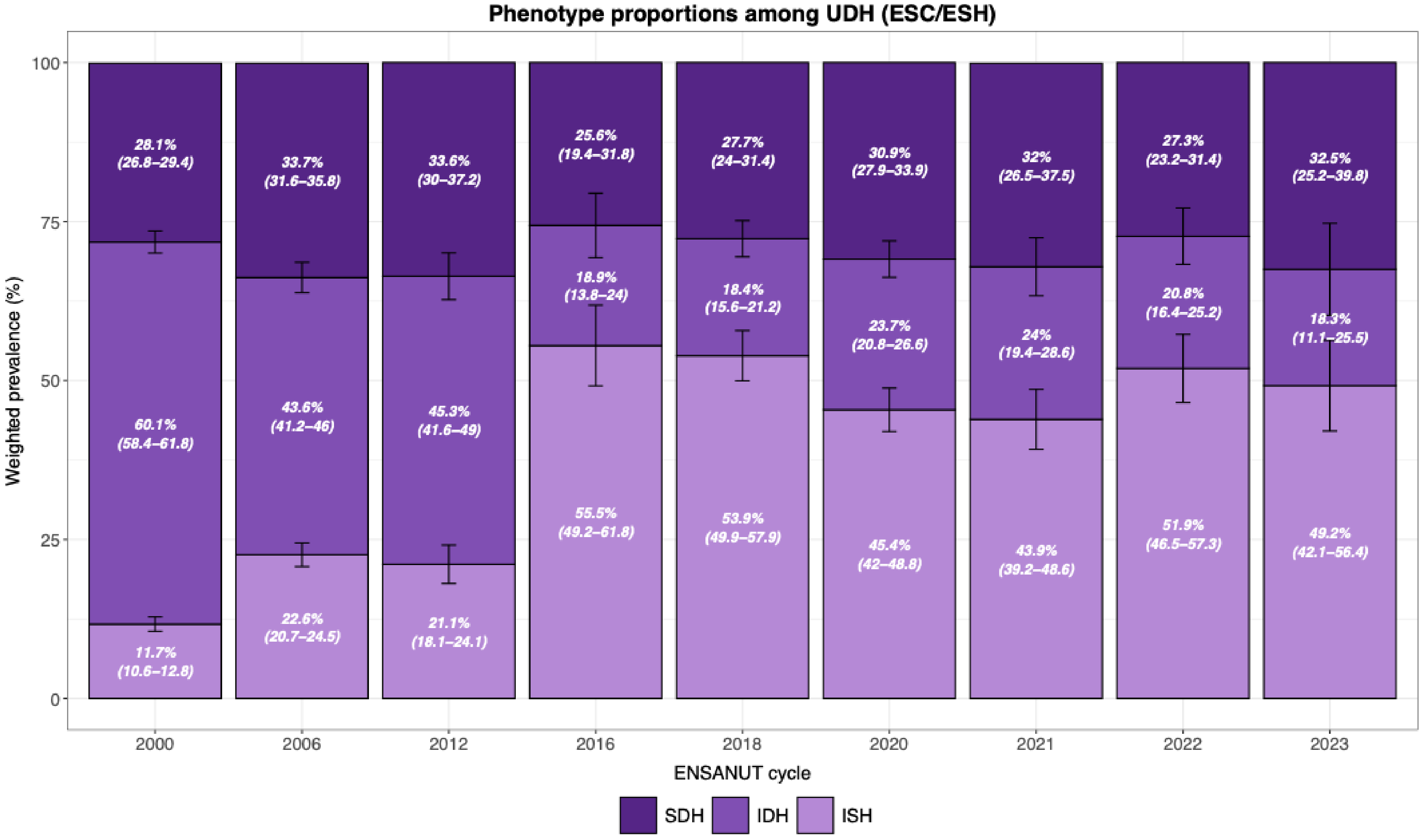

**Supplementary Figure 5. Change in the prevalence of isolated systolic hypertension (ISH) in Mexico**. Trends in ISH based on ENSANUT 2000-2023 data, stratified by (A) age group, (B) sex, (C) body mass index (BMI)-defined obesity, and (D) diabetes status.

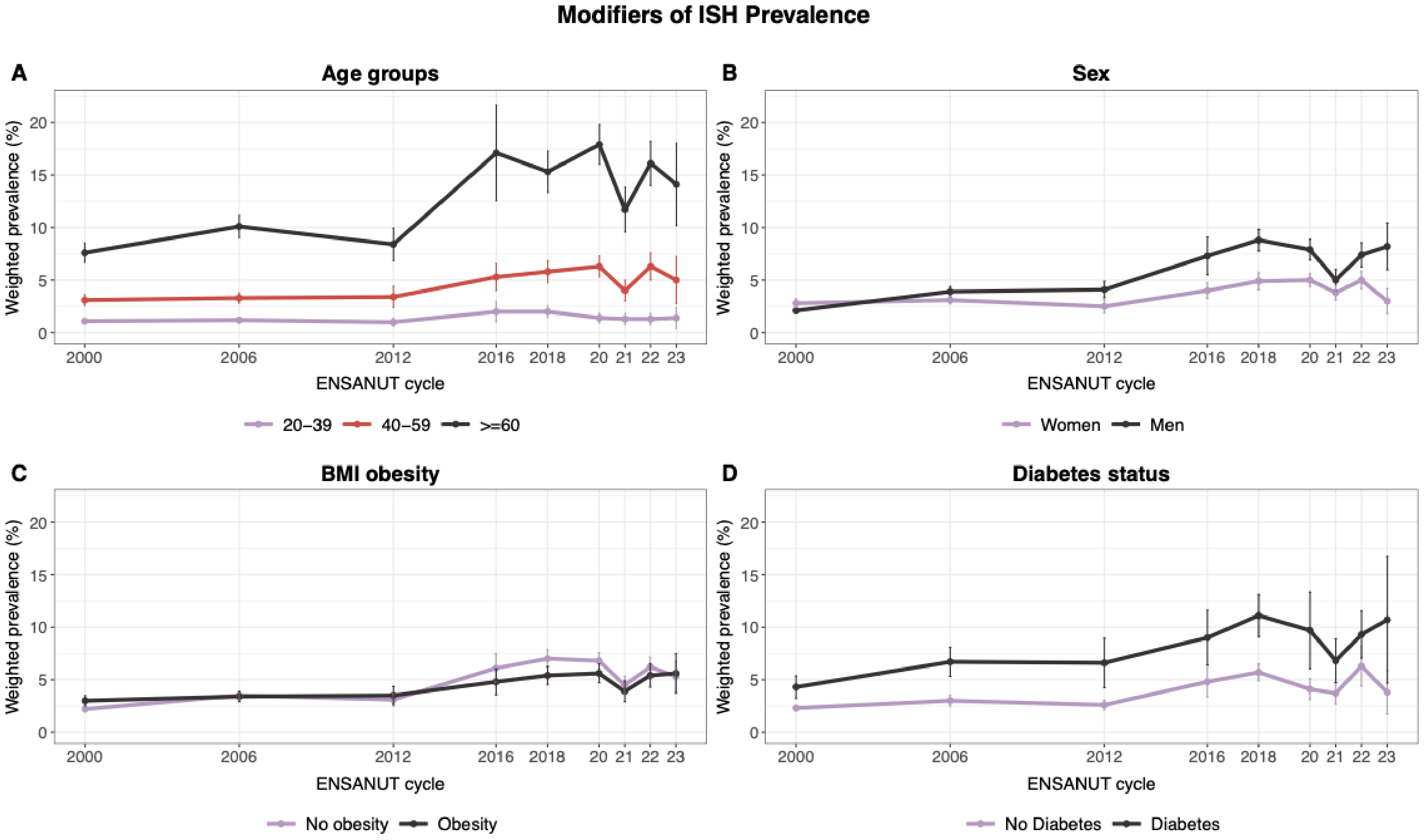

**<u>Supplementary Figure 6</u>. Change in the prevalence of isolated diastolic hypertension (IDH) in Mexico**. Trends in IDH based on ENSANUT 2000-2023 data, stratified by (A) age group, (B) sex, (C) body mass index (BMI)-defined obesity, and (D) diabetes status.

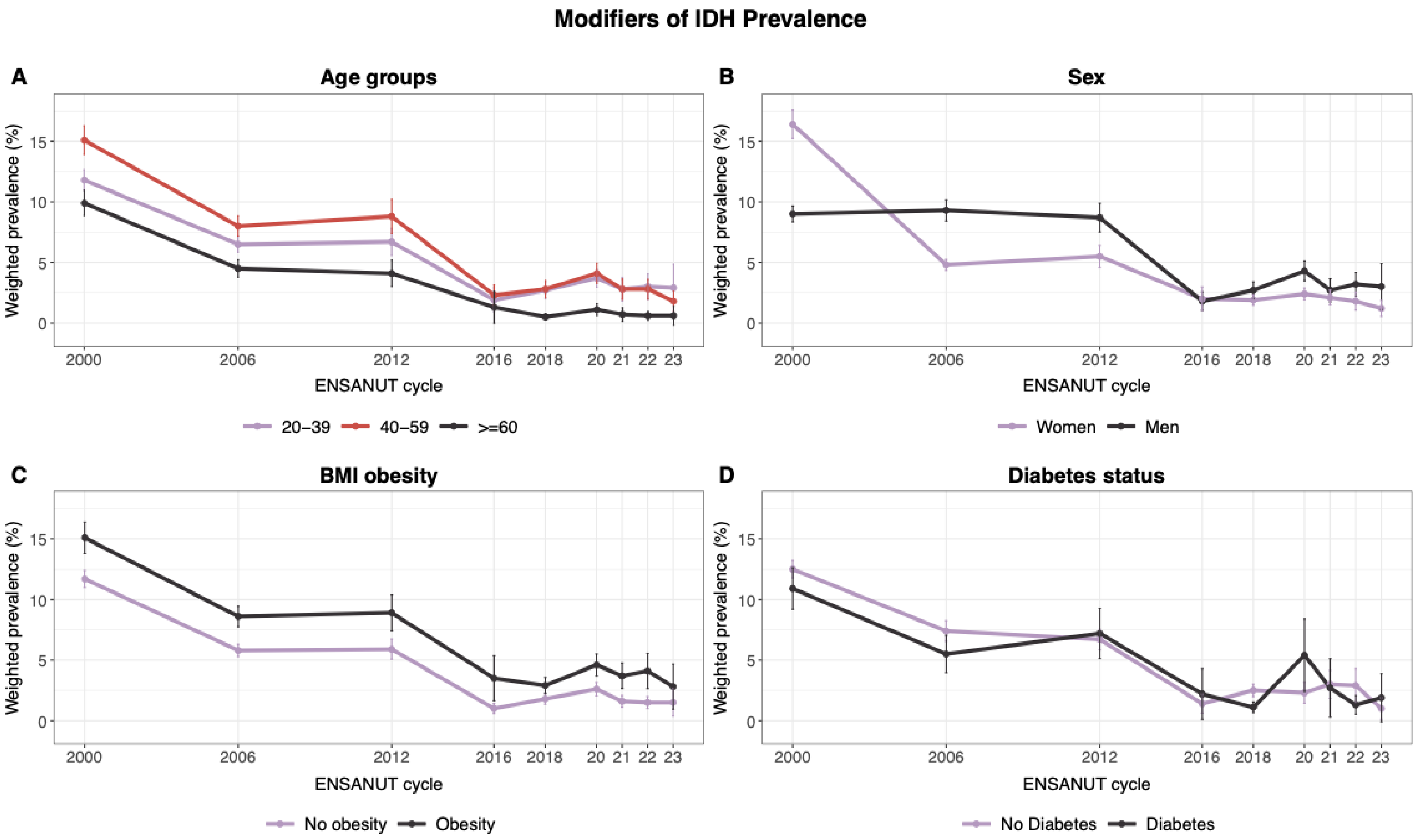

**<u>Supplementary Figure 7</u>. Change in the prevalence of systolic-diastolic hypertension (SDH) in Mexico**. Trends in SDH based on ENSANUT 2000-2023 data, stratified by (A) age group, (B) sex, (C) body mass index (BMI)-defined obesity, and (D) diabetes status.

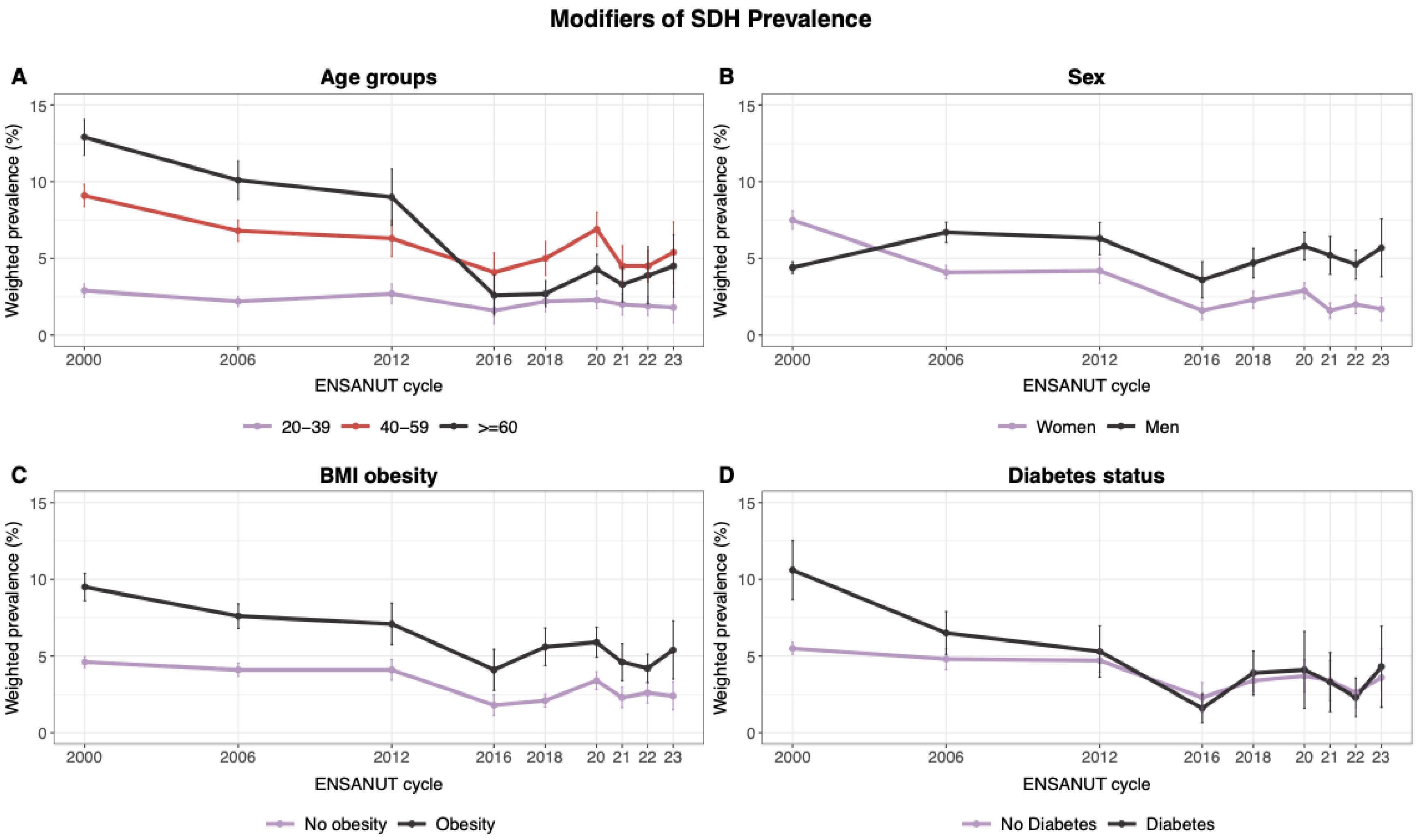
